# Transcriptomic Profiling of the Amygdala of Children with Autism Spectrum Disorder

**DOI:** 10.64898/2026.03.04.26347554

**Authors:** Jobin Babu, Ayushi Lal, Lavanya Challagundla, Obie Allen, Madeline Griffin, Barbara Gisabella, Harry Pantazopoulos

## Abstract

A growing number of studies point to a key role of the amygdala in Autism Spectrum Disorders (ASD). The amygdala is involved in several processes in ASD including emotional valence, facial recognition, regulation of social learning, empathy, and anxiety. Brain imaging and postmortem studies demonstrate altered amygdala development in children with ASD, associated with impairment in social behavior and anxiety. There is limited information regarding the molecular pathology of the amygdala in children with ASD. We conducted RNAseq profiling on postmortem amygdala samples from male children (4-14 yrs old) with ASD (n=8) and normotypic male children (n=6). Furthermore, we conducted drug repurposing analysis to identify compounds predicted to reverse the transcriptomic signatures identified in order to identify potential therapeutic targets for development of early intervention treatments. Full transcriptome gene expression profiling implicated molecular pathways involved in neuroimmune signaling, glycogen and carbohydrate metabolism, matrix metalloproteases, neurodevelopment, estrogen receptor signaling, and synaptic signaling. Targeted pathway analysis of the top 10% of differentially expressed genes implicated pathways involved in extracellular matrix organization, immune signaling, and synaptic signaling. Our drug repurposing analysis identified sleep modifying compounds and anti-inflammatory compounds including COX2 and GSK3 inhibitors amongst the top predicted therapeutic compound classifications. PDGF receptor tyrosine kinase inhibitors were identified as a top potential therapeutic mechanism of action. Our results point to alterations in immune signaling, extracellular matrix organization, and synaptic signaling in the amygdala of children with ASD. Furthermore, our results identified a number of potential therapeutic drug targets for development of early intervention strategies.

## Introduction

Autism Spectrum Disorder (ASD) is a neurodevelopmental disorder characterized by social and communication impairments, along with stereotypic behaviors, that is typically diagnosed during childhood ^1–3^. In recent decades, the diagnosis of ASD has substantially increased in the United States, from one in 44 children in 2018 to one in 36 in 2020 ^4, 5^, highlighting a need for improved understanding of the neurobiological processes underlying ASD that can serve as targets for development of new treatments. Several lines of evidence point to the amygdala as a key brain region in the neurocircuitry processes involved in ASD. The amygdala is critically involved in several processes implicated in ASD including the processing of emotional valence of environmental stimuli, as well as the regulation of social learning, empathy, and anxiety ^6–10^. In particular, the amygdala is involved in facial recognition and direct eye contact ^11–13^, processes that are often impaired in people with ASD ^14–16^. Studies have demonstrated reduced amygdala activity while viewing faces in people with ASD compared to normotypic control subjects, which provides further support for an underlying amygdala pathology in this process ^17^. Case studies from normotypic subjects with amygdala damage compared with subjects with ASD provide additional support for a core amygdala pathology in ASD ^18^.

Several studies suggest neurodevelopmental dysfunction of the amygdala in ASD. For example, brain imaging studies demonstrate that the amygdala has a prolonged postnatal developmental course compared to other brain areas, and that this postnatal development is sex specific, suggesting a role for hormonal factors ^19, 20^. In addition, a number of studies report that the volume of the amygdala is larger in children wih ASD ^21–23^, along with volume changes in brain regions connected to the amygdala ^24, 25^. These changes are found to be most prominent in children with more severe social impairment ^24, 25^. Furthermore, differences in amygdala volume are associated with greater levels of anxiety in children with ASD ^26^.

Human postmortem studies have identified alterations in neuron numbers and dendritic spines in the amygdala that display an altered developmental trajectory in people with ASD ^27, 28^. While the number of neurons increases from childhood to adulthood in the typical developing human amygdala, children with ASD display greater neuron numbers at an early age compared with age matched control subjects, with an altered developmental trajectory resulting in decreased numbers in in adulthood compared with control subjects ^27^. Similarly, children with ASD display greater numbers of dendritic spines in the amygdala compared to age matched normotypic controls, and an altered developmental trajectory resulting in decreased amygdala dendritic spine numbers in adulthood ^28^. Collectively, these studies provide high resolution neuroanatomical support for the volumetric MRI evidence of altered amygdala development in ASD.

Despite the growing evidence for altered neurodevelopment of the amygdala in children with ASD, there is limited information regarding the underlying molecular pathways in this region in children with ASD ^29^, which limits the development of early intervention therapeutic strategies. Our recent study of the hippocampus of children with ASD utilizing a largely overlapping cohort to the current study, identified an increased expression of molecules involved in neuroimmune signaling and extracellular matrix organization (ECM), along with a decreased expression of molecules involved in synaptic signaling ^30^. Abnormalities in neurodevelopmental and neuroimmune processes are key features of ASD ^27, 28^. In addition, extracellular matrix molecules (ECM), including chondroitin sulphate proteoglycans (CSPGs) and their endogenous proteases, are critically involved in mediating immune responses and represent key factors at the intersection of neuroimmune signaling, neurodevelopment, and synaptic regulation ^31–36^. The amygdala is highly interconnected with the hippocampus ^37, 38^, and these two regions work together during the storage and retrieval of emotional memories ^39^, suggesting potential overlap in the altered molecular processes during brain development in children with ASD.

Several studies indicate that early diagnosis of ASD can have a significant positive impact on therapeutic efficacy and quality of life ^40, 41^, providing potential opportunities for early intervention strategies. Therefore, identifying neurobiological processes involved during early ages can guide development of early intervention strategies. We used a cohort of human postmortem amygdala samples from male children with ASD and age matched normotypic control subjects (4-14 yrs old) (Table 1) to examine the molecular pathways altered in the amygdala of children with ASD. We focused on a neurodevelopmental time-window encompassing stages of synaptic development and refinement that represents an age range for early intervention. Furthermore, we used iLINCS drug repurposing analysis on our transcriptomic data in order to identify top predicted therapeutic mechanisms of action and compounds, and in order to test for predicted relevance of compounds currently used for ASD treatment and/or proposed as potential ASD treatments.

**Table 1:** Subjects and demographic information.

| Age | Sex | Race | RIN | PMI<br>(hrs) | Hemisphere | Epilepsy | Cause of<br>Death |
| --- | --- | --- | --- | --- | --- | --- | --- |
| 1-5 | Male | Black | 7.4 | 18 | Left | No | natural |
| 11-15 | Male | Black | 6.8 | 16 | Left | No | natural |
| 11-15 | Male | Black | 7.5 | 11 | Left | No | accidental |
| 11-15 | Male | Black | 6.8 | 20 | Left | No | undetermined |
| 11-15 | Male | unknown | 2.4 | 15 | Left | No | suicide |
| 6-10 | Male | Black | 7.6 | 16 | Left | No | accidental |
| mean<br>± SD |  |  |  |  |  |  |  |
| 10.3±<br>3.8 | 6 M | 5B, 1U | 6.4±<br>2.0 | 16.0<br>±3.0 | 6 Left | 6 No |  |
| 6-10 | Male | N/A | 7.1 | 12 | Left | N/A | N/A |
| 11-15 | Male | Hispanic | 7.1 | 27 | Left | No | natural |
| 11-15 | Male | White | 6.5 | 9 | Left | No | accidental |
| 6-10 | Male | White | 7.7 | 3 | Left | No | natural |
| 1-5 | Male | White | 6.0 | 21 | Left | No | accidental |
| 11-15 | Male | N/A | 4.8 | 20 | Left | N/A | natural |
| 11-15 | Male | Black | 8.3 | 22 | Left | Yes | natural |
| 11-15 | Male | N/A | 6.5 | 15 | Left | N/A | suicide |
| mean<br>± SD |  |  |  |  |  |  |  |
| 9.9±3.<br>3 | 8 M | 1B, 3W,<br>1H, 3<br>N/A | 6.8±<br>1.1 | 16.1<br>±7.9 | 8 Left | 4 No, 1<br>Yes, 3<br>N/A |  |

## Methods

### Human Subjects

Postmortem human brain amygdala samples of ASD (n=8) and non-ASD (n=6) male children (4-14 yrs old, Table 1) were obtained from the NIH NeuroBioBank at the University of Maryland, Baltimore MD which obtained informed consent. IRB approval was obtained from the Univ. of Maryland IRB committee. Cohort size was determined based on previous studies ^29, 30, 42^. Frozen blocks were sectioned at 30 µm thickness using a Leica CM 3050 S cryostat (Leica Biosystems, Buffalo Grove, IL). This study is limited to males only, as ASD is four times more prevalent in males than females ^43^, and to avoid any additional sex and hormonal variabilities.

### RNAseq & Alignment

RNA isolation, library preparation, and next generation sequencing was performed by the Molecular and Genomics Core Facility at the University of Mississippi Medical Center, as described previously ^44^. Total RNA was isolated from tissue samples using the Invitrogen PureLink RNA Mini kit with Trizol (Life Technologies; Carlsbad, CA, USA) following manufacturer protocol. Quality control of total RNA was assessed using the Agilent TapeStation for quality and Qubit Fluorometer for concentration measures. Libraries were prepared using the TruSeq Stranded mRNA Library Prep Kit per manufacturer’s protocol using on average 161 ng of RNA per sample. Libraries were index-tagged, pooled for multiplexing, and then sequencing was performed on the Illumina NextSeq 2000 platform using a paired-end P4 200 cycle with ∼1.113 billion reads (%QC30=91.9) being generated. FastQC (v0.11.9; Andrews S, 2010) was used to assess raw read quality and MultiQC (v1.12; Ewels et al., 2016) summarized preprocessing metrics. The reads were then quantified using Salmon (v1.10.0; Patro et al., 2017) against the Ensembl GRCh38 release 114 reference transcriptome for *Homo sapiens*. Transcript annotations were obtained from the corresponding Ensembl GTF file. Transcript-to-gene mappings were generated from the GTF annotation using the Bioconductor (v3.21) packages GenomicFeatures (v1.60.0) and txdbmaker (v1.4.2) (Lawrence et al., 2013; Pagès et al., 2025). Transcript-level estimates were imported into R (v4.5) and summarized to gene-level counts using tximport (v1.36.1; Soneson et al.,2016).

### RNAseq Differential Expression Analysis

Differential expression (DE) of genes (i.e. mRNA and non-coding RNA) was assessed between subjects with ASD and controls using DESeq2 R Package (v1.48.2; Love et al., 2014). Prior to DE analysis, low-count genes were removed by retaining only those genes with at least 10 counts in a minimum of 3 samples. The control group was set as the reference level for all comparisons. A gene was considered differentially expressed if the p-value did not exceed 0.05. Heatmap visualizations of the top 30 differentially expressed genes (ranked by p-value) were generated using normalized counts scaled by row, with genes (rows) hierarchically clustered and samples (columns) ordered by group, using the pheatmap R package (v1.0.13; Kolde 2015). Volcano plots were generated using ggplot2 (v4.0.0; Wickham 2016).

### Gene Set Enrichment Analysis (GSEA)

GSEA was conducted using the clusterProfiler (v4.16.0; Yu et al., 2012) and ReactomePA (v1.52.0; Yu & He, 2016) R packages. The gseGO and gsePathway functions were used for GO Biological Process and Reactome pathway gene set collections, respectively. For both analyses, genes were ranked by log2 fold change as the ranking metric. Genes were then analyzed using default permutation procedures with parameters minGSSize = 20, maxGSSize = 500, and pAdjustMethod = "none” with post hoc filtering at p < 0.05. Normalized enrichment scores (NES) were used to determine directionality of pathway regulation. Leading edge gene analysis was performed using the GSEA GO Biological Process results, where core enrichment genes were extracted from each significant pathway and pooled per direction. Gene frequency was counted as the number of significant pathways in which each gene appeared, with the top 20 genes per direction ranked by descending frequency identified as the most consistently enriched leading edge genes.

### Targeted Pathway Analysis

Targeted pathway analysis was performed on disease gene sets composed of the top and bottom 10% of genes ranked by log2 fold change (greatest absolute log2FC) using the enrichR R package (v3.4; Kuleshov et al., 2016). The following databases were queried: GO Biological Process 2025, GO Cellular Component 2025, GO Molecular Function 2025, and Reactome Pathways 2024. Gene symbols were used as input after conversion from Ensembl IDs using the GTF annotation file. Pathways with P-value < 0.05 were considered significant. Results were ranked and visualized using the enrichR combined score, which is computed as the product of the log-transformed p-value and the z-score deviation from the expected rank, normalized per database (Chen et al., 2013). Raw sequencing reads were deposited in the NIH Data Archive Collection (accession pending). All differential expression results are available in Supplementary Data.

### Identification of perturbagens altering gene expression

The Library of Integrated Network-Based Cellular Signatures (LINCS) is a National Institutes of Health initiative designed to map molecular responses to environmental and internal stressors (Pilarczyk et al., 2022). The project employs the L1000 assay, a gene expression profiling platform that measures 978 landmark genes to generate transcriptomic signatures, capturing approximately 82% of the transcriptome’s information content (Subramanian et al., 2017). For this study, genes identified as differentially expressed (p < 0.05) in the ASD versus control analysis were uploaded programmatically to the iLINCS environment through the SignatureMeta API. These signatures were then compared against the L1000 Chemical Perturbagens collection (LIB_5) using the findConcordances function (Pilarczyk et al., 2022). The platform quantified similarity between the disease profile and reference perturbation signatures using a weighted correlation of log fold-change vectors, where weights were derived from the negative log10 p-values associated with both datasets (Koleti et al., 2018). Perturbagens with connectivity scores ≥ 0.321 were classified as concordant, whereas those with scores ≤ −0.321 were considered discordant. Mechanism of action annotations were retrieved from the iLINCS CompoundMOA API, and drug approval status was obtained from the ChEMBL API (www.ebi.ac.uk/chembl), with compounds classified as Approved (Phase 4), Investigational (Phase 2-3), or Experimental (Phase 0-1) (Zdrazil et al., 2024; Davies et al., 2015). For analysis of proposed ASD therapeutic compounds, perturbagens with discordant scores were classified into psychiatric medications, sleep modulating compounds, hormone signaling compounds, anti-inflammatory and immune signaling compounds, GSK inhibitors, calcium channel blockers and extracellular matrix proteases.

## Results

### Differential Gene Expression Analysis

Hierarchical cluster analysis of the top 30 DEGs revealed co-expression patterns among genes, with samples grouped by diagnosis showing the expected separation between ASD and control, with inter-subject variability (Fig. 1A). RNAseq gene expression profiling identified 542 differentially expressed genes in the amygdala of children with ASD, including genes implicated in estrogen signaling (ESR1, EGR1, AKR1C3, PRLR), neurodevelopment (NEUROG2, SOX11, RUNX1, EMX2), neuroimmune signaling (CXCL10, PLA2G5, HLA-B, HLA-DRB1), sleep and circadian rhythms (NPAS4, ADRB1, PPARGC1B), synaptic signaling (GABBR1, HTR1A, GRM1, FOSB) metabolism (ATP2A1, IGFL2, SAT1), and ECM organization (MMP16, MMP2, COL4A4, ADAMTSL1, ADAMTS17), (Fig. 1B, STable 1).

**Figure 1:**
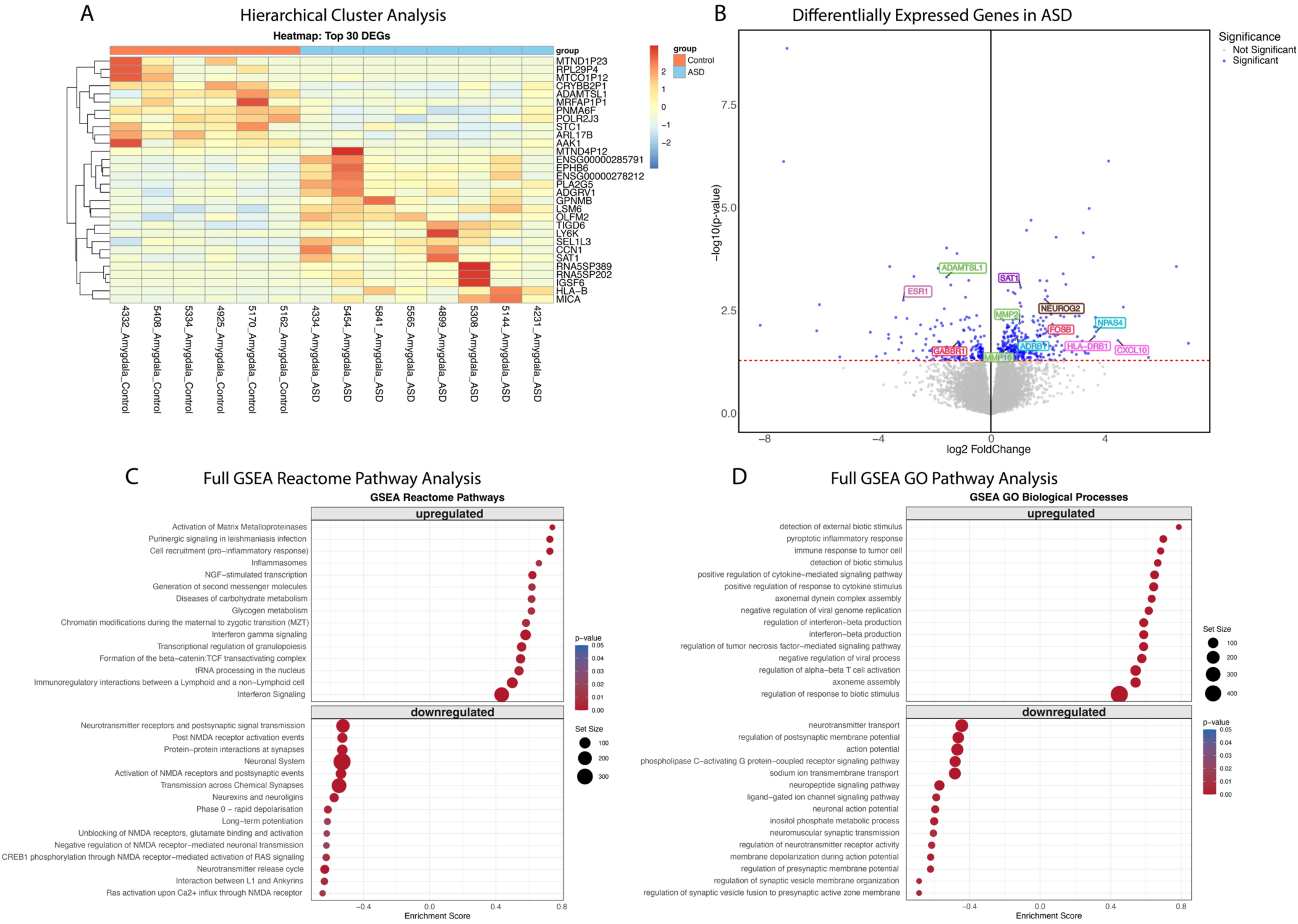
Differential Gene Expression Pathways in the Amygdala of Children with ASD. (A) Heatmap of the top 30 genes differentially expressed between 8 ASD and 6 non-ASD control subjects in the amygdala (p < 0.05), generated from normalized counts scaled by row with genes hierarchically clustered and samples ordered by group. (B) Volcano plot of DGE: ASD vs. non-ASD controls. On the x-axis is log fold change of genes in subjects with ASD compared to controls, points to the right of 0 represent genes that are increased, and points to the left of 0 genes that are decreased, in ASD compared to controls. Statistical significance is displayed on the y-axis, and p < 0.05 are labeled in blue. Select genes with both high fold change and significance are labeled. Full transcriptome Reactome pathway analysis of amygdala pathways of subjects with ASD including top 15 upregulated pathways and downregulated pathways (C). (D) Full transcriptome GO pathway analysis of amygdala pathways of subjects with ASD including top 15 upregulated pathways and downregulated pathways. X-axis represents the normalized enrichment score (NES). This score considers the overrepresentation of a gene set in a ranked list of all genes between two conditions (i.e., ASD vs. control). A positive NES indicates significant upregulation at the top of the list, a negative NES signifies significant down-regulation at the bottom, while an NES close to 0 suggests no enrichment. Larger absolute values of NES imply stronger enrichment.

### Pathway Analysis

Pathway analysis (Figure 1, Tables S2&3) conducted using GSEA takes advantage of information on continuous expression changes from all transcribed genes to determine the biological processes (gene sets) that are statistically significantly different between the disorder and control groups ^45^. Full transcriptomic GSEA Reactome pathway analysis identified upregulation of pathways involved in activation of matrix metalloproteinases (R-HSA-1592389), inflammasomes (R-HSA-622312), diseases of carbohydrate metabolism (R-HSA-5663084), glycogen metabolism (R-HSA-8982491), and interferon signaling (R-HSA-913531) (Fig. 1C, Table S3). Similarly, full GSEA GO pathway analysis identified upregulated pyroptotic inflammatory response (GO:0070269), positive-regulation of cytokine mediated signaling (GO:0001961), regulation of alpha-beta T-cell activation (GO:0046634), axoneme assembly (GO:0035082), and regulation of response to biotic stimulus (GO:0002831) (Fig. 1D, Table S3).

In contrast, full GSEA Reactome pathway analysis identified downregulation of several pathways involved in synaptic signaling and synaptic transmission including transmission across the chemical synapse (R-HSA-112315), neuronal systems (R-HSA-112316), long-term potentiation (R-HSA-9620244), neurotransmitter release cycle (R-HSA-112310), and interaction between L1 and ankyrins (R-HSA-445095) (Fig. 1C, Table S2). Full GSEA GO pathway analysis identified similar downregulation in synaptic signaling pathways including neurotransmitter transport (GO:0006836), action potential (GO:0001508), regulation of neurotransmitter receptor activity (GO:0010469), and regulation of synaptic vesicle membrane organization (GO:1901632) (Fig. 1D, Table S3).

Reactome tree plot of the full pathway GSEA analysis identified additional pathways including several neurodevelopmental processes and epigenetic regulation processes, estrogen receptor signaling, collagen biosynthesis (Fig. 2A). Similarly, a tree plot of the GO full GSEA pathway analysis identified several processes including positive regulation of phosphorylation, negative regulation of cell adhesion, sex differences, neural precursor cell proliferation, cerebral cortex development, and axon guidance in addition to a number of immune signaling pathways (Fig. 2B), also highlighted in the full GSEA enrichment plot (Fig. 2C).

**Figure 2:**
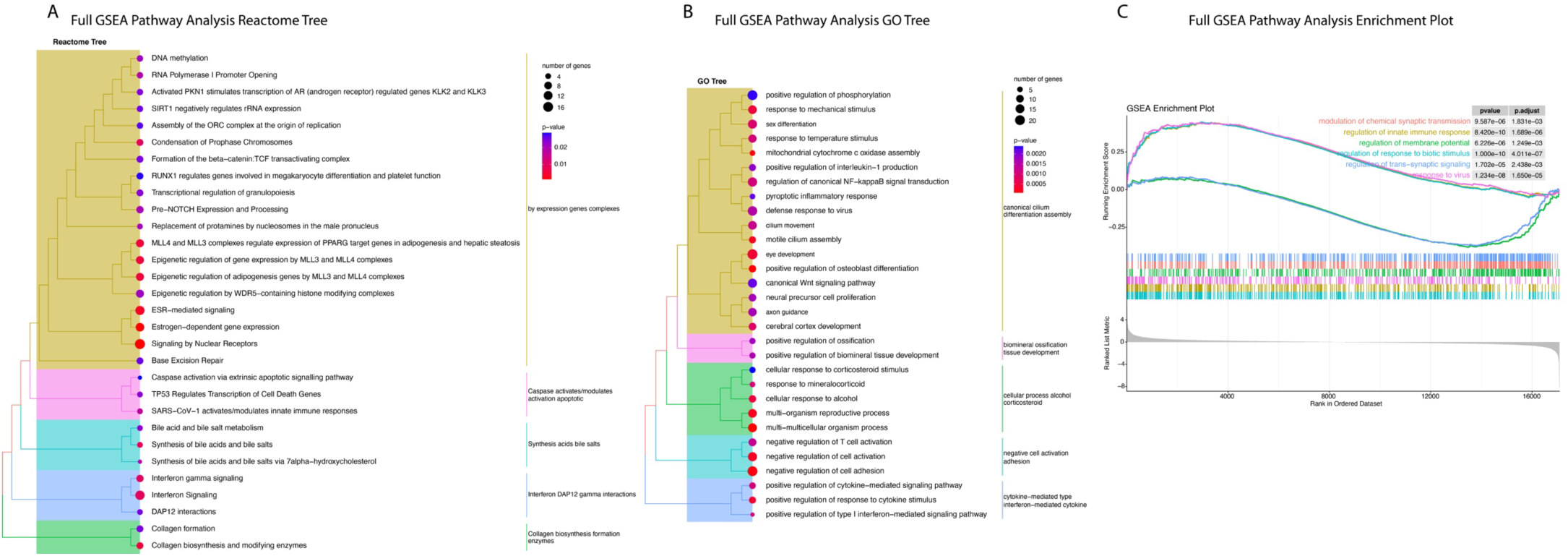
Full Transcriptome Reactome and GO Tree and Enrichment Plots. Reactome tree (A) and GO tree (B) plots of pathways altered in the amygdala of children with ASD, with pathways hierarchically clustered by similarity and colored by p-value. (C) GSEA enrichment plot of the upregulated and downregulated Gene Ontology (GO) Biological Process terms. Genes were ranked by log2 fold change comparing children with ASD to non-ASD controls.

### EnrichR Pathway Analysis

Targeted pathway analysis of the top and bottom 10% differentially expressed genes implicated upregulation of several GO Biological Process pathways involved in immune signaling including negative regulation of lymphocyte proliferation (GO:0050672), regulation of T-cell proliferation (GO: 0042129), and positive regulation of cytokine production (GO:0001818) along with upregulation of pathways involved in ECM regulation including extracellular matrix organization (GO:0030198) and extracellular structure organization (GO: 0043062) (Fig. 3A, STable 6), (Fig. 3A). Downregulated GO Biological Process pathways identified several pathways involved in synaptic transmission including chemical synaptic transmission (GO:0007268), synaptic transmission, GABAergic (GO0051932), anterograde trans-synaptic signaling (GO:0098916), and neurotransmitter transport (GO0006836), (Fig. 3A, STable 7).

**Figure 3:**
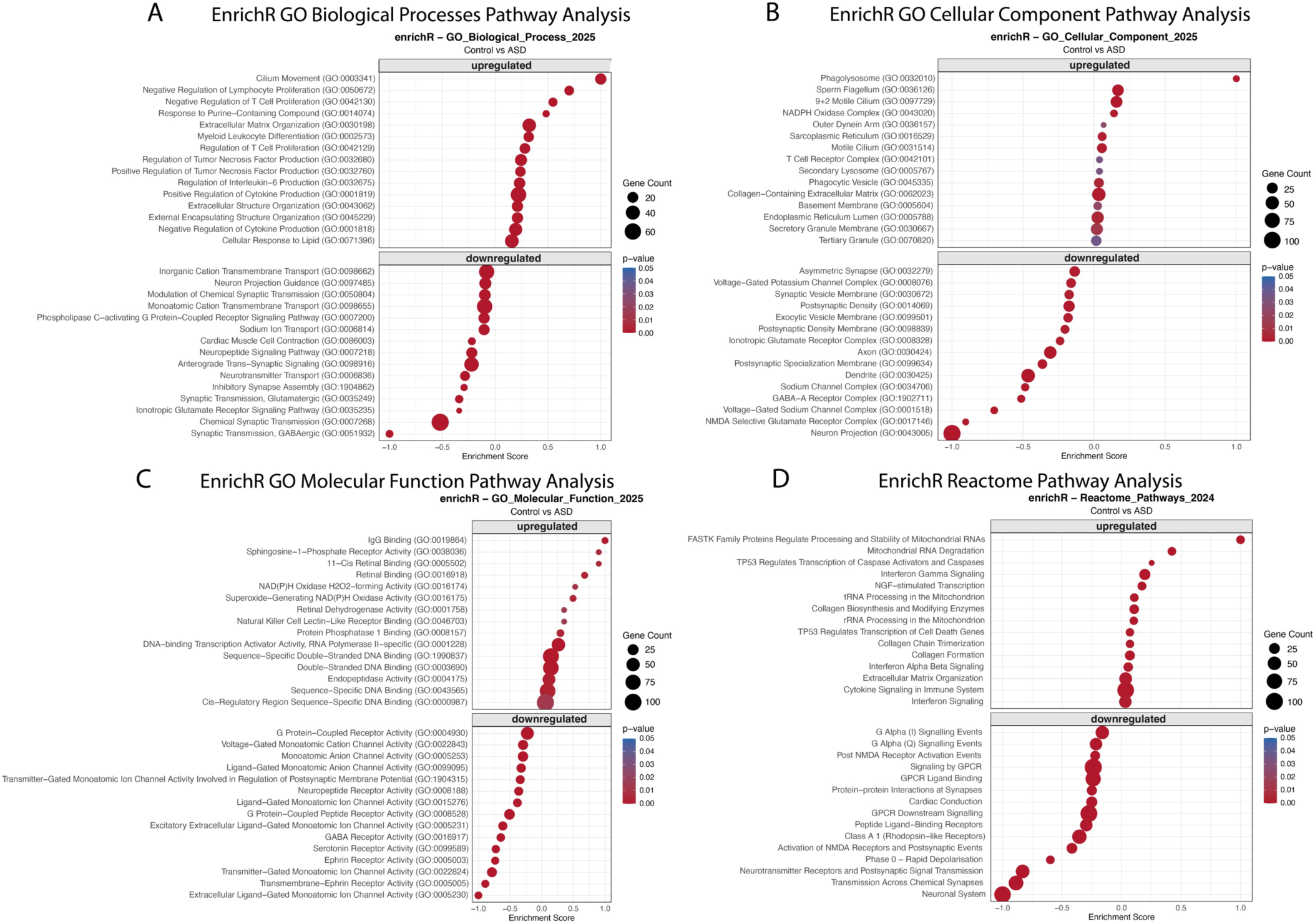
EnrichR Analysis. Targeted pathway analysis of the top and bottom 10% of genes ranked by log2 fold change, showing the top 15 upregulated and downregulated terms for (A) GO Biological Process, (B) GO Cellular Component, (C) GO Molecular Function, and (D) Reactome pathways. The x-axis represents the enrichR combined score normalized within each direction.

Targeted pathway analysis of GO cellular component pathways implicated upregulation of several components involved in immune response including phagolysosome (GO: 0032010), 9+2 motile cilium (GO: 0097729), and secretory granule membrane (GO: 0030667), along with the extracellular matrix component collagen-containing extracellular matrix (GO: 0062023), (Fig. 3B, STable 8). Downregulated cellular component pathways included a range of synaptic signaling and neurodevelopmental components such as neuronal projection (GO: 0043005), GABA-A receptor complex (GO: 1902711), dendrite (GO: 0030425), axon (GO: 0030424), and postsynaptic specialization membrane (GO: 0099634), (Fig. 3B, STable 9).

Similarly, GO Molecular Function targeted pathway analysis identified upregulation in a number of pathways involved in immune signaling including IgG binding (GO: 0019864) and natural killer cell lectin-like receptor binding (0046703) in addition to several DNA regulatory pathways including cis-regulatory region sequence-specific DNA binding pathway (GO:0000987), sequence-specific double-stranded DNA binding (GO: 1990837), and double-stranded DNA binding (GO: 0003690), (Fig. 3C, STable 10). Downregulated molecular function pathways included GABA receptor activity (GO: 0016917), serotonin receptor activity (GO: 0099589), and extracellular ligand-gated monoatomic ion channel activity (GO: 0005230), (Fig. 3C, STable 11).

Targeted Reactome pathway analysis identified upregulation of several ECM pathways including extracellular matrix organization, collagen formation, and collagen biosynthesis and modifying enzymes, along with several immune signaling pathways (Fig. 3D, STable 12). Downregulated targeted Reactome pathways largely consisted of pathways involved in synaptic transmission such as neuronal systems, transmission across chemical synapses, activation of NMDA receptors and postsynaptic events, and GPCR ligand binding (Fig. 3D, STable 13).

### Leading edge Gene Analysis

Leading edge gene analysis identified the genes that are most influential for enrichment of significant pathways (Figure 4, Tables S4&5)). Leading edge genes are identified based on the frequency with which they are identified in biological pathways; their expression is not necessarily statistically significant in disease compared to control. The top 20 upregulated leading edge genes derived from our GSEA analysis consisted largely of immune signaling genes including TGFß1, LGALS9, HLA-DRB1, TLR4 and IL1B (Fig. 4A, STable 4). In comparison, the top 20 downregulated leading edge genes consisted of a number of genes involved in synaptic regulation and extracellular matrix organization such as NRXN1, NLGN1, STXBP1, RELN, and genes involved in neurodevelopment such as the transcription factor MEF2C and PRNP (Fig. 4B, STable 5).

**Figure 4:**
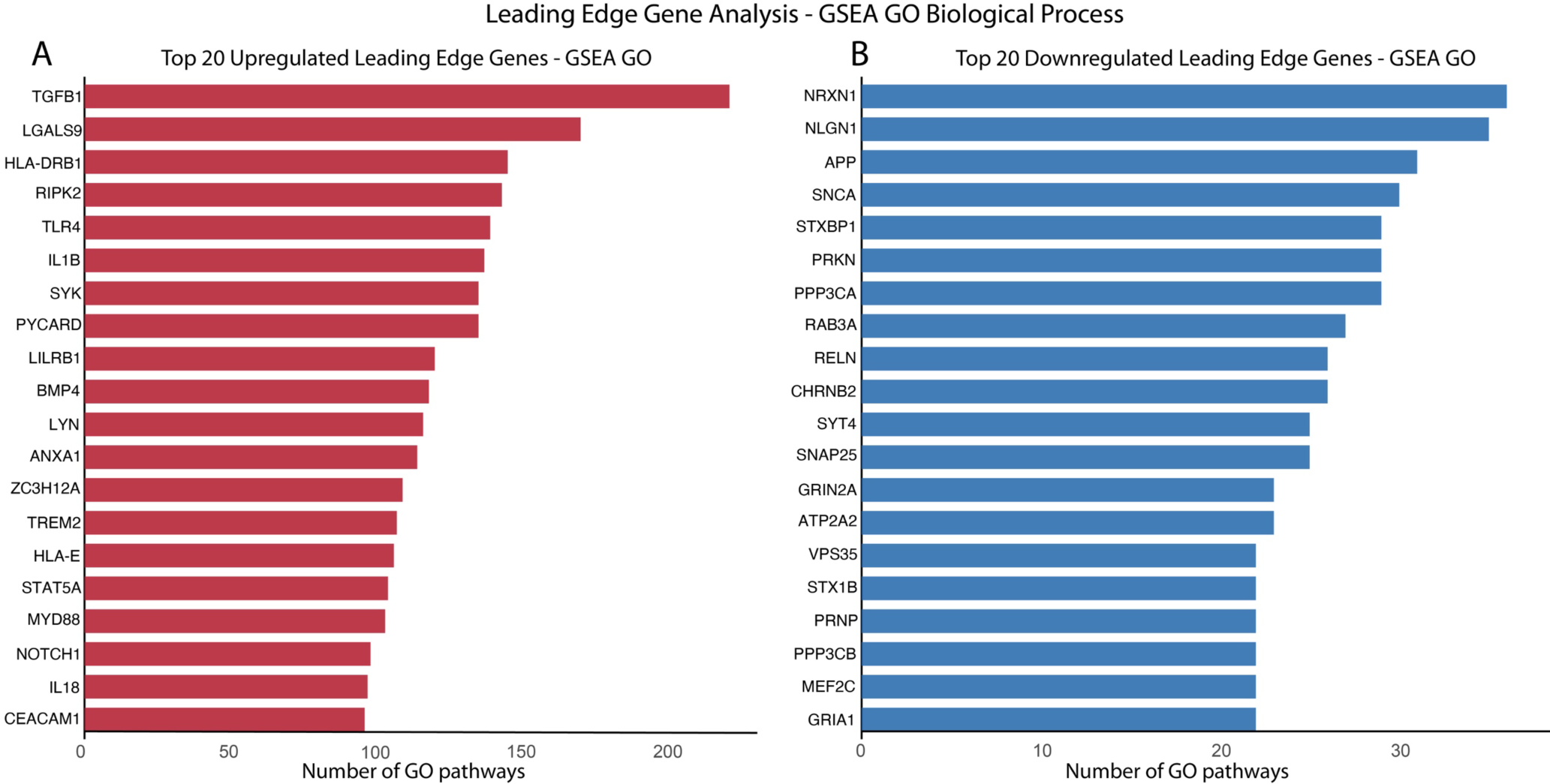
Leading-Edge Gene Analysis. Top 20 upregulated (A) and downregulated (B) leading-edge genes in the amygdala of subjects with ASD. X-axis represents the number of GO pathways involved.

### iLINCS Drug Repurposing Analysis

Signature-based connectivity analysis utilizing the Library of Integrated Network-based Signatures (LINCS) database examined chemical perturbagens that were dissimilar (discordant) or similar (concordant) to the transcriptional signatures of ASD (Figure 5A&B, STables 16-21). The signature reversion principle suggests that chemical perturbagens that are discordant with the disease signature may induce gene expression changes that “reverse” disease-associated gene expression signatures ^46^. Equally, concordant chemical perturbagen signatures may indicate drugs that induce gene expression changes similar to those found in the disease state, informing on the underlying gene targets that may be implicated in disease.

**Figure 5:**
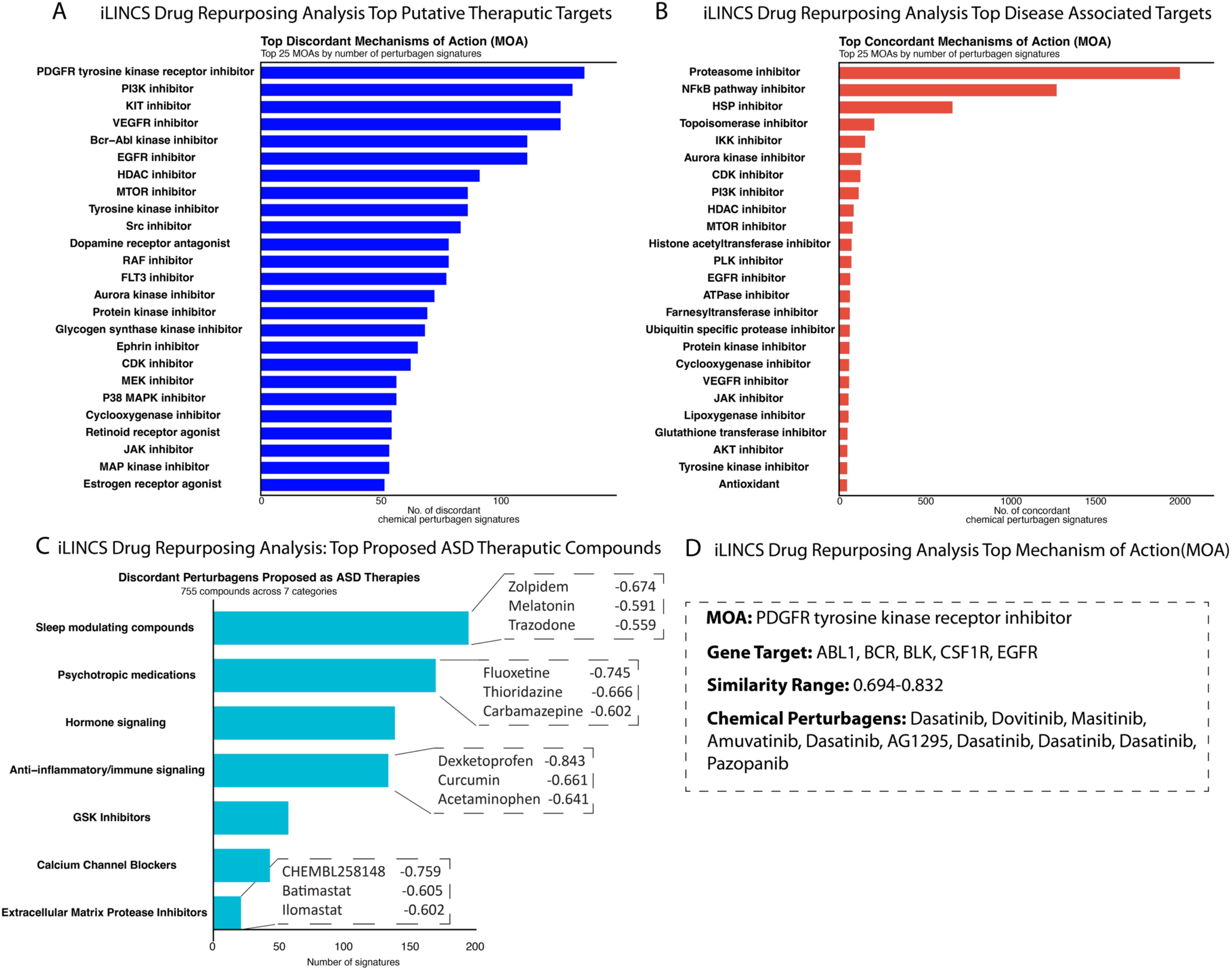
iLINCS Drug Repurposing Analysis. (A) Top 25 discordant and (B) concordant chemical perturbagens identified through iLINCS perturbagen analysis. The x-axis represents the number of chemical signatures associated with a mechanism of action (MOA). (C) Discordant drug classes used for or under investigation as ASD treatment targets, arranged by number of associated chemical signatures. (D) Top iLINCS discordant mechanisms of action with corresponding gene targets, similarity score range, and associated chemical perturbagens.

The iLINCS analysis identified 7808 concordant (i.e., perturbagens that may induce gene expression changes) and 6529 discordant (putative therapeutic) mechanisms of action (MOAs).

The top discordant mechanisms of action (MOAs) identified included PDGF receptor tyrosine kinase inhibitors, PI3K inhibitors, KIT inhibitors, VEGFR inhibitors, Glycogen synthase kinase (GSK) inhibitors, and cyclooxygenase (COX) inhibitors (Fig. 5A, STable 18). The overall top MOA identified was PDGF receptor tyrosine kinase inhibitor, with a similarity score range of 0.694-0.832 encompassing several compounds including Dasatinib, Dovotinib, and Masitinib and several gene targets including ABL1, CBR, CSF1R and EGFR (Fig. 4D).

Top concordant pathways, i.e. putative causative MOAs, included Proteasome inhibitors, NFkB pathway inhibitors, Topoisomerase inhibitors, PI3K inhibitors, HDAC inhibitors, ATPase inhibitors and AKT inhibitors (Fig. 5B, STable 19). Our analysis of potential ASD therapeutic compounds classified into categories for compounds either used to treat ASD symptoms or under investigation for potential ASD treatment identified sleep modulating compounds as the top potential therapeutic drug class, which included sleep aides such as zolpidem, melatonin, and trazodone (Fig. 5C, STable 20). This was followed by psychiatric drugs composed of antipsychotics, antidepressants, and anticonvulsants used to treat ASD symptoms and proposed ASD treatment targets, which included Fluoxetine, Thioridazine, and Carbamazepine (Fig. 4C, STables 20&21). Anti-inflammatory and immune signaling compounds including COX inhibitors such as Dexketoprofen and Acetaminophen, and natural compounds such as Curcumin also displayed high similarity scores as potential therapeutic compounds, with Dexketoprofen displaying the highest similarity score across all compounds (−0.843) (Fig. 5C, STables 20&21). A smaller number of GSK inhibitors and matrix metalloprotease inhibitors, under investigation as potential ASD therapeutic targets, also displayed high similarity scores (Fig. 4C, STables 20&21).

## Discussion

Our results add to the growing evidence implicating altered amygdala neurodevelopment in ASD and represent to our knowledge the first combined transcriptomic profiling and drug repurposing study of the amygdala of children with ASD. Our findings provide a molecular profile of the amygdala in ASD during a critical postnatal developmental window that has been implicated by prior volumetric and neuroanatomical studies ^21–28^. Our findings of estrogen receptor signaling pathways (Fig. 2), together with evidence that postnatal development of the amygdala is sex specific ^19, 20^, suggest a potential contribution of hormonal factors in amygdala neurodevelopmental alterations that may contribute to the male prevalence of ASD ^43^. Our leading edge gene analysis identified several genes implicated in prior ASD studies invluding NRXN1, NLGN1, TGFB1, ANK3 and SHANK2 ^47–52^, as well as genes implicated in a study of the hippocampus of a largely overlapping cohort (MMP9, IL1B, MEF2C, and CACNA1C) ^30^. Furthermore, our drug repurposing analysis provides a molecular link to several antipsychotics, antidepressants, and anti-convulsant drugs used to treat ASD and identifies a number of potential therapeutic compounds and drug classes that may serve as promising early intervention targets.

### Neuroimmune Signaling and Synaptic Signaling

Our findings of altered neuroimmune signaling in the amygdala of children with ASD add to the growing evidence for the involvement of neuroimmune processes in ASD ^53–56^ and link these processes to a key neurodevelopmental window of this brain region implicated by brain imaging and neuroanatomical studies ^21–28^. Upregulation of immune signaling pathways was one of the most consistent results across our full GSEA and targets pathway analyses, suggesting a critical involvement of enhanced immune signaling during postnatal development of the amygdala. Interferon gamma signaling and T-cell activation were some of the top immune pathways implicated, along with cellular components including phagolysosome, cilium motility, phagocytic vesicles, and T-cell receptor complexes, together indicating active neuroimmune response. Furthermore, upregulated leading edge genes largely consisted of genes involved in neuroimmune processes including TGFß1, LGALS9, HLA-DRB1, TLR4 and ILB1 (Fig 4A, STable 4). ILB1 mRNA and protein levels were upregulated in the hippocampus in a largely overlapping cohort ^30^, suggesting shared processes between these brain regions in children with ASD. Taken together with upregulated expression of microglial activation genes such as HLA-DRB1 and matrix metalloproteases including MMP2 and MMP16, this suggests possible enhanced microglial pruning of synapses during this neurodevelopmental period. This is in line with prior neuroanatomical evidence of decreased dendritic spines in the amygdala of subjects with ASD from childhood to adulthood in comparison to increased dendritic spines in this developmental period in control subjects ^28^.

Furthermore, our results displaying downregulation of synaptic pathways are one of our most consistent findings and provides further support for this enhanced synaptic pruning during amygdala postnatal development in children with ASD. Neuroimmune molecules are critically involved in the regulation of synaptic plasticity and neurodevelopmental processes ^57^, and increased cytokine levels during early developmental stages are associated with increased risk of developing ASD ^58, 59^. Enhanced neuroimmune signaling therefore may underlie synaptic decreases and changes in amygdala volume reported in ASD, potentially contributing to social impairment and anxiety in children with ASD ^24, 25^ ^26^. Identification of anti-inflammatory and immune signaling compounds as top predicted therapeutic targets for our observed transcriptomic changes in the amygdala of children with ASD (Fig. 5) provides futher support for the key role of immune signaling and suggests that anti-inflammatory compounds may be a promising early intervention strategy for postnatal amygdala neurodevelopmental alterations. In particular, COX inhibitors including Dexketoprofen and Acetaminophen displayed some of the strongest predictive (similarity) scores of all potential therapeutic compounds.

### Extracellular Matrix Molecules

Our full and targeted pathway analyses identified several ECM pathways upregulated in the amygdala of children with ASD, in line with our previous findings in the hippocampus of the same subjects ^30^. ECM molecules are critically involved in the regulation of neurodevelopment, synaptic regulation, and neuroimmune processes and represent key factors that may mediate these processes in ASD ^31–36^. In particular, we observed an upregulation of matrix metalloproteinase pathways, including increased expression of MMP16 and MMP2. Furthermore, several ECM proteases including MMP9 and MMP2 were identified as upregulated leading edge genes (Fig.4, Stable 4). MMP9 mRNA and protein levels were also upregulated in the hippocampus of a largely overlapping cohort ^30^, indicating shared processes across this brain circuit during development. MMPs are primarily produced by glial cells including microglia ^60, 61^ and contribute to remodeling of synaptic ECM structures ^62^. MMP16 is highly expressed in younger microglial cells and associated with synaptic remodeling ^63^. Our observed upregulation of microglial activation genes such as HLA-DRB1, together with upregulated matrix metalloprotease pathways indicates enhanced microglial pruning of synapses during amygdala postnatal development in children with ASD and suggests activated neuroimmune signaling may contribute to our observed ECM alterations. However, genome-wide association studies (GWAS) have reported several ECM genes associated with ASD, including genes encoding for endogenous proteases ^64–68^, suggesting that ECM alterations may be due in part to genetic factors. In either case, upregulated activation of matrix metalloproteases and ECM organization pathways may impair perisynaptic ECM and perineuronal nets (PNNs), which are critical for the stabilization of synapses ^62, 69–76^. Impairment of these ECM synaptic stabilizing structures may in turn contribute to the broad decreases of synaptic signaling pathways observed in the amygdala of children with ASD.

Evidence from animal models supports the hypothesis that increased MMP expression during development contributes to decreased PNNs and synaptic destabilization ^70–72^. Fragile-X syndrome is a monogenetic disease in which approximately 30% of patients display symptoms of ASD. Several animal models of Fragile-X syndrome, a monogenetic disease associated with ASD, display increased MMP expression and reduced synaptic ECM structures in the amygdala and hippocampus, along with impaired fear memory ^69–72^. Pharmacological inhibition or genetic knockdown of MMPs during development restores ECM structures and reduces anxiety ^70, 71^, suggesting that MMP inhibition during this postnatal developmental window may prevent neurodevelopmental alterations that may contribute to aspects of ASD. Our drug reproposing analysis identified a number of MMP inhibitors and cathepsin ECM protease inhibitors, including Batimastat and Ilomastat, providing evidence from brain samples of children with ASD that supports preclinical studies suggesting that MMP inhibitors may be a promising early intervention target for ASD.

Upregulated ECM pathways also included several pathways involved in collagen biosynthesis and remodeling. ECMs, including collagen molecules, are involved in blood-brain barrier (BBB) regulation ^77^, suggesting that these processes may be involved in altered BBB composition in the amygdala of children with ASD, in agreement with prior reports of BBB dysfunction in ASD ^78, 79^. Disrupted BBB composition may contribute to neuroimmune dysfunction in ASD, which could subsequently impact synaptic regulation, highlighting the role of ECM molecules in many neurobiological processes implicated in ASD.

### Potential Therapeutic and Causative Targets

Our drug repurposing analysis identified a number of potential therapeutic targets including targets that are currently under investigation for development of ASD therapies such as PDGF receptor tyrosine kinase inhibitors, GSK inhibitors, matrix metalloproteinase inhibitors, and anti-inflammatory compounds. These results provide a key link between preclinical drug development studies and brain imaging studies that consistently implicate altered postnatal amygdala development associated with social impairment and anxiety in ASD ^21–28^.

Previous studies have implicated PDGFRA signaling in ASD ^80, 81^. For example, increased gene expression of PDGFRA and associated immune signaling pathways was implicated in the cerebellum of subjects with ASD and in brain samples from offspring of mice from mothers with maternal immune activation, indicating that enhanced PDGF signaling is associated with neuroimmune activation in ASD ^80^. Furthermore, increased serum levels of PDGF have been reported in male children with ASD ^81^. PDGFR tyrosine kinase receptor inhibitors such as Dasatinib, and another of our top targets, PI3K inhibitors, are also potent inhibitors of the microglial activating factor CSF1R ^82–84^, indicating that this top target may alleviate microglial mediated neuroimmune activation in ASD. Along this line, we identified anti-inflammatory and immune signaling compounds as potential therapeutic targets. This includes a number of COX inhibitors such as Dexketoprofen and Acetaminophen. with Dexketoprofen, which displayed the strongest similarity score across all compounds examined, has also been reported to have anti-epileptic effects in preclinical studies ^85^, and epilepsy is a common comorbid condition in children with ASD.

Glycogen synthase kinase (GSK) inhibitors have been a focus of several preclinical studies and clinical trials for ASD ^86–88^. Preclinical models have demonstrated increased GSK signaling in transgenic mouse models associated with ASD, and efficacy for GSK inhibitors in alleviating aberrant protein synthesis along with learning and memory deficits ^87^, brain connectivity and dendritic spine densities ^86^. The GSK3ß inhibitor Tideglusib is currently being investigated in a clinical trial on adolescent subjects with ASD (ClinicalTrials.gov ID: NCT02586935). Our findings provide support for the use of GSK inhibitors as early intervention treatment for ASD and suggest these compounds may alleviate alterations in the amygdala during postnatal development.

Psychotropic medications have long been used in children with ASD to treat symptoms, albeit with limited efficacy ^89–93^. We identified psychotropic medications as top predicted therapeutic drug classes (Fig. 5C). This includes antipsychotics such as thioridazine and clozapine, antidepressants such as escitalopram and fluoxetine and trazodone, and anticonvulsant medications including carbamazepine. Our findings provide molecular evidence suggesting that these medications may be partially effective in reversing gene expression changes occurring in the amygdala during postnatal development in children with ASD.

Sleep modulating compounds were the top predicted therapeutic drug class from our analysis of drug classes currently used or under investigation for ASD treatment (Fig. 5C). Furthermore, several genes involved in sleep and circadian rhythm regulation were implicated as upregulated leading edge genes, including ADA, NR1D1, and ADORA2A (STable 4). Growing evidence suggests that sleep disruption is a risk factor for ASD ^94–99^.. Sleep is at the intersection of several processes consistently implicated in ASD, including neurodevelopment, neuroimmune signaling, and synaptic regulation ^94^. Furthermore, sleep disturbances are common in people with ASD, with up to 86% of children and adolescence reporting sleep problems ^95–97^. Polysomnography studies demonstrate that insomnia, restless sleep, and circadian rhythm disturbances are the most prevalent sleep problems ^98^. Recent findings indicate that male children age 8-14 years old with insufficient sleep display a greater risk for developing ASD ^99^. In addition, preclinical studies suggest that early life sleep disruption may be a causative factor for neurodevelopmental alterations associated with ASD. Early life sleep disruption in prairie voles results in a number of male specific behavioral and neurocircuitry alterations in adults reflective of ASD including deficits in pair bond formation and novel object recognition ^100^. Furthermore, adult voles with early life sleep disruption display increased cortical dendritic spines, impaired cognitive flexibility ^101^ and reduced REM sleep ^102^. Our findings provide further support that disrupted sleep may contribute to neurodevelopmental alterations in male children with ASD and suggest sleep modulating compounds may be a promising early intervention strategy. Several compounds identified in our study have been proposed for the treatment of children with ASD including melatonin ^103–105^, flumazenil ^106^, clonidine ^107^, and the orexin antagonist SB408124 ^108^.

Our analysis also identified several potential factors that can contribute to the changes we observed in the amygdala of children with ASD. Proteosome inhibitors displayed the greatest number of concordant chemical perturbagen signatures, suggesting this is a top potential causative factor for the transcriptional alterations in the amygdala of children with ASD (Fig. 5B). Proteosome dysregulation has been implicated in a number of studies of ASD ^109^. Fmr1 mouse models display a significant increase of protein degradation through the ubiquitin-proteasome system, and the protease inhibitor bortezomib alleviates the molecular and behavioral alterations in these mice ^109^. This is in contrast to our drug repurposing analysis identifying proteasome inhibitors as the top potential causative factor for alterations in the amygdala, suggesting that proteasome inhibitors may be effective broadly as a treatment for ASD but may not be ideal for amygdala processes during this age range in male children with ASD. Taken together, these findings suggest that alteration of proteasome inhibition can potentially contribute to transcriptomic changes occurring in the amygdala during this developmental window, and this may be dependent on the dose of the protease inhibitors and the age, tissue and species. A number of studies have consistently reported that prenatal exposure to the HDAC and NFkB inhibitor valproic acid results in increased risk of developing ASD ^110–112^. HDAC inhibitors and NFkB inhibitors were identified as top potential causative factors in our study, suggesting that this potential risk may also apply during postnatal ages.

### Technical Considerations

Our study consisted of bulk amygdala gene expression analyses, which does not allow for analysis of expression changes in specific amygdala nuclei or anterior to posterior gradients in expression. Furthermore, our bulk amygdala transcriptomics profiling approach did not allow for evaluation of cell type specific changes or confirmation with protein expression. Future studies consisting of amygdala subregion specific profiling together with protein analysis and larger numbers of subjects may provide greater information regarding the amygdala neurocircuitry alterations in children with ASD. Our drug repurposing analysis is an approach to identify potential therapeutic candidates and potential causative mechanisms of action but is a correlational analysis limited to the cell lines and drug classes available in the iLINCS database, as well as to our amygdala transcriptomic dataset. Future studies following up on specific drug targets are needed to confirm and develop these targets as potential therapeutic or causative compounds.

In summary, our results identify transcriptomic alterations in the amygdala of children with ASD during a key neurodevelopmental period. These findings indicate neuroimmune signaling, synaptic signaling, and ECM abnormalities may contribute to altered postnatal development of the amygdala in ASD. Furthermore, our results identified a number of potential therapeutic compounds including sleep modulating compounds, anti-inflammatory compounds, and PDFG tyrosine kinase receptor inhibitors that may serve as targets for development of early intervention therapies.

## Supporting information

Babu et al Supplemental Tables

## Acknowledgments

We thank the NIH NeuroBioBank (https://neurobiobank.nih.gov/) for making the human brain samples available. The authors deeply appreciate the invaluable contributions made by the families consenting to donate brain tissue. This research was supported by the Baszucki Brain Research Foundation to HP; Joe W. and Dorothy Dorsett Brown Foundation Neuroscience Initiative: AWD-001292, NIGMS P20-GM144041 to BG. The work performed through the UMMC Molecular and Genomics Facility is supported, in part, by funds from the NIGMS, including Molecular Center of Health and Disease (P20GM144041) and Mississippi INBRE (P20GM103476).

## Data Availability Statement

Gene expression profiling data will be publicly available on NIMH Data Archive upon manuscript publication. All other data are available in the main text or supplementary materials.

## Author Contributions Statement

HP and JB designed the studies, analyzed data, and wrote the manuscript. BG, AY, and LC contributed to study design, data interpretation, and manuscript preparation. OA contributed to data collection. MG contributed manuscript preparation. All authors contributed to the article and approved the submitted version.

## References

1. Lai MC, Lombardo MV, Baron-Cohen S. Autism. Lancet. 2014;383(9920):896–910. Epub 2013/10/01. doi: 10.1016/S0140-6736(13)61539-1. PubMed PMID: 24074734.

2. Prevention CfDCa. CDC estimates 1 in 59 children has been identified with autism spectrum disorder;2018.

3. Schendel DE, Thorsteinsson E. Cumulative Incidence of Autism Into Adulthood for Birth Cohorts in Denmark, 1980-2012. Jama. 2018;320(17):1811–3. doi: 10.1001/jama.2018.11328. PubMed PMID: 30398592; PMCID: PMC6248103.

4. Maenner MJ, Warren Z, Williams AR, Amoakohene E, Bakian AV, Bilder DA, Durkin MS, Fitzgerald RT, Furnier SM, Hughes MM, Ladd-Acosta CM, McArthur D, Pas ET, Salinas A, Vehorn A, Williams S, Esler A, Grzybowski A, Hall-Lande J, Nguyen RHN, Pierce K, Zahorodny W, Hudson A, Hallas L, Mancilla KC, Patrick M, Shenouda J, Sidwell K, DiRienzo M, Gutierrez J, Spivey MH, Lopez M, Pettygrove S, Schwenk YD, Washington A, Shaw KA. Prevalence and Characteristics of Autism Spectrum Disorder Among Children Aged 8 Years - Autism and Developmental Disabilities Monitoring Network, 11 Sites, United States, 2020. MMWR Surveill Summ. 2023;72(2):1–14. Epub 20230324. doi: 10.15585/mmwr.ss7202a1. PubMed PMID: 36952288; PMCID: PMC10042614.

5. Maenner MJ, Shaw KA, Bakian AV, Bilder DA, Durkin MS, Esler A, Furnier SM, Hallas L, Hall-Lande J, Hudson A, Hughes MM, Patrick M, Pierce K, Poynter JN, Salinas A, Shenouda J, Vehorn A, Warren Z, Constantino JN, DiRienzo M, Fitzgerald RT, Grzybowski A, Spivey MH, Pettygrove S, Zahorodny W, Ali A, Andrews JG, Baroud T, Gutierrez J, Hewitt A, Lee LC, Lopez M, Mancilla KC, McArthur D, Schwenk YD, Washington A, Williams S, Cogswell ME. Prevalence and Characteristics of Autism Spectrum Disorder Among Children Aged 8 Years - Autism and Developmental Disabilities Monitoring Network, 11 Sites, United States, 2018. MMWR Surveill Summ. 2021;70(11):1–16. Epub 20211203. doi: 10.15585/mmwr.ss7011a1. PubMed PMID: 34855725; PMCID: PMC8639024.

6. Baron-Cohen S, Ring HA, Wheelwright S, Bullmore ET, Brammer MJ, Simmons A, Williams SC. Social intelligence in the normal and autistic brain: an fMRI study. Eur J Neurosci. 1999;11(6):1891–8. Epub 1999/05/21. doi: 10.1046/j.1460-9568.1999.00621.x. PubMed PMID: 10336657.

7. Meyer-Lindenberg H, Moessnang C, Oakley B, Ahmad J, Mason L, Jones EJH, Hayward HL, Cooke J, Crawley D, Holt R, Tillmann J, Charman T, Baron-Cohen S, Banaschewski T, Beckmann C, Tost H, Meyer-Lindenberg A, Buitelaar JK, Murphy DG, Brammer MJ, Loth E. Facial expression recognition is linked to clinical and neurofunctional differences in autism. Mol Autism. 2022;13(1):43. Epub 2022/11/12. doi: 10.1186/s13229-022-00520-7. PubMed PMID: 36357905; PMCID: PMC9650909.

8. Kleinhans NM, Richards T, Weaver K, Johnson LC, Greenson J, Dawson G, Aylward E. Association between amygdala response to emotional faces and social anxiety in autism spectrum disorders. Neuropsychologia. 2010;48(12):3665–70. Epub 2010/07/27. doi: 10.1016/j.neuropsychologia.2010.07.022. PubMed PMID: 20655320; PMCID: PMC3426451.

9. Yeung M, Engin E, Treit D. Anxiolytic-like effects of somatostatin isoforms SST 14 and SST 28 in two animal models (Rattus norvegicus) after intra-amygdalar and intra-septal microinfusions. Psychopharmacology (Berl). 2011;216(4):557–67. Epub 2011/03/23. doi: 10.1007/s00213-011-2248-x. PubMed PMID: 21424237.

10. Hurlemann R, Patin A, Onur OA, Cohen MX, Baumgartner T, Metzler S, Dziobek I, Gallinat J, Wagner M, Maier W, Kendrick KM. Oxytocin enhances amygdala-dependent, socially reinforced learning and emotional empathy in humans. J Neurosci. 2010;30(14):4999–5007. doi: 10.1523/JNEUROSCI.5538-09.2010. PubMed PMID: 20371820; PMCID: PMC6632777.

11. Johnson MH. Subcortical face processing. Nat Rev Neurosci. 2005;6(10):766–74. doi: 10.1038/nrn1766. PubMed PMID: 16276354.

12. Skuse D. Fear Recognition and the Neural Basis of Social Cognition. Child Adolesc Ment Health. 2003;8(2):50–60. doi: 10.1111/1475-3588.00047. PubMed PMID: 32797554.

13. Wang S, Yu R, Tyszka JM, Zhen S, Kovach C, Sun S, Huang Y, Hurlemann R, Ross IB, Chung JM, Mamelak AN, Adolphs R, Rutishauser U. The human amygdala parametrically encodes the intensity of specific facial emotions and their categorical ambiguity. Nature communications. 2017;8:14821. Epub 20170421. doi: 10.1038/ncomms14821. PubMed PMID: 28429707; PMCID: PMC5413952.

14. Papagiannopoulou EA, Chitty KM, Hermens DF, Hickie IB, Lagopoulos J. A systematic review and meta-analysis of eye-tracking studies in children with autism spectrum disorders. Social neuroscience. 2014;9(6):610–32. Epub 20140702. doi: 10.1080/17470919.2014.934966. PubMed PMID: 24988218.

15. Pelphrey KA, Sasson NJ, Reznick JS, Paul G, Goldman BD, Piven J. Visual scanning of faces in autism. Journal of autism and developmental disorders. 2002;32(4):249–61. doi: 10.1023/a:1016374617369. PubMed PMID: 12199131.

16. Griffin JW, Bauer R, Scherf KS. A quantitative meta-analysis of face recognition deficits in autism: 40 years of research. Psychological bulletin. 2021;147(3):268–92. Epub 20201026. doi: 10.1037/bul0000310. PubMed PMID: 33104376; PMCID: PMC8961473.

17. Costa C, Cristea IA, Dal Bo E, Melloni C, Gentili C. Brain activity during facial processing in autism spectrum disorder: an activation likelihood estimation (ALE) meta-analysis of neuroimaging studies. Journal of child psychology and psychiatry, and allied disciplines. 2021;62(12):1412–24. Epub 20210315. doi: 10.1111/jcpp.13412. PubMed PMID: 33723876.

18. Sweeten TL, Posey DJ, Shekhar A, McDougle CJ. The amygdala and related structures in the pathophysiology of autism. Pharmacol Biochem Behav. 2002;71(3):449–55. doi: 10.1016/s0091-3057(01)00697-9. PubMed PMID: 11830179.

19. Giedd JN, Castellanos FX, Rajapakse JC, Vaituzis AC, Rapoport JL. Sexual dimorphism of the developing human brain. Prog Neuropsychopharmacol Biol Psychiatry. 1997;21(8):1185–201. doi: 10.1016/s0278-5846(97)00158-9. PubMed PMID: 9460086.

20. Giedd JN, Vaituzis AC, Hamburger SD, Lange N, Rajapakse JC, Kaysen D, Vauss YC, Rapoport JL. Quantitative MRI of the temporal lobe, amygdala, and hippocampus in normal human development: ages 4-18 years. J Comp Neurol. 1996;366(2):223–30. doi: 10.1002/(SICI)1096-9861(19960304)366:2<223::AID-CNE3>3.0.CO;2-7. PubMed PMID: 8698883.

21. Schumann CM, Hamstra J, Goodlin-Jones BL, Lotspeich LJ, Kwon H, Buonocore MH, Lammers CR, Reiss AL, Amaral DG. The amygdala is enlarged in children but not adolescents with autism; the hippocampus is enlarged at all ages. J Neurosci. 2004;24(28):6392–401. doi: 10.1523/JNEUROSCI.1297-04.2004. PubMed PMID: 15254095; PMCID: PMC6729537.

22. Mosconi MW, Cody-Hazlett H, Poe MD, Gerig G, Gimpel-Smith R, Piven J. Longitudinal study of amygdala volume and joint attention in 2- to 4-year-old children with autism. Arch Gen Psychiatry. 2009;66(5):509–16. doi: 10.1001/archgenpsychiatry.2009.19. PubMed PMID: 19414710; PMCID: PMC3156446.

23. Nordahl CW, Scholz R, Yang X, Buonocore MH, Simon T, Rogers S, Amaral DG. Increased rate of amygdala growth in children aged 2 to 4 years with autism spectrum disorders: a longitudinal study. Arch Gen Psychiatry. 2012;69(1):53–61. doi: 10.1001/archgenpsychiatry.2011.145. PubMed PMID: 22213789; PMCID: PMC3632313.

24. Lee JK, Andrews DS, Ozturk A, Solomon M, Rogers S, Amaral DG, Nordahl CW. Altered Development of Amygdala-Connected Brain Regions in Males and Females with Autism. J Neurosci. 2022;42(31):6145–55. Epub 20220627. doi: 10.1523/JNEUROSCI.0053-22.2022. PubMed PMID: 35760533; PMCID: PMC9351637.

25. Schumann CM, Barnes CC, Lord C, Courchesne E. Amygdala enlargement in toddlers with autism related to severity of social and communication impairments. Biol Psychiatry. 2009;66(10):942–9. Epub 20090902. doi: 10.1016/j.biopsych.2009.07.007. PubMed PMID: 19726029; PMCID: PMC2795360.

26. Andrews DS, Aksman L, Kerns CM, Lee JK, Winder-Patel BM, Harvey DJ, Waizbard-Bartov E, Heath B, Solomon M, Rogers SJ, Altmann A, Nordahl CW, Amaral DG. Association of Amygdala Development With Different Forms of Anxiety in Autism Spectrum Disorder. Biol Psychiatry. 2022;91(11):977–87. Epub 20220202. doi: 10.1016/j.biopsych.2022.01.016. PubMed PMID: 35341582; PMCID: PMC9116934.

27. Avino TA, Barger N, Vargas MV, Carlson EL, Amaral DG, Bauman MD, Schumann CM. Neuron numbers increase in the human amygdala from birth to adulthood, but not in autism. Proc Natl Acad Sci U S A. 2018;115(14):3710–5. Epub 2018/03/22. doi: 10.1073/pnas.1801912115. PubMed PMID: 29559529; PMCID: PMC5889677.

28. Weir RK, Bauman MD, Jacobs B, Schumann CM. Protracted dendritic growth in the typically developing human amygdala and increased spine density in young ASD brains. J Comp Neurol. 2018;526(2):262–74. Epub 2017/09/21. doi: 10.1002/cne.24332. PubMed PMID: 28929566; PMCID: PMC5728110.

29. Sorrells SF, Paredes MF, Velmeshev D, Herranz-Perez V, Sandoval K, Mayer S, Chang EF, Insausti R, Kriegstein AR, Rubenstein JL, Manuel Garcia-Verdugo J, Huang EJ, Alvarez-Buylla A. Immature excitatory neurons develop during adolescence in the human amygdala. Nature communications. 2019;10(1):2748. Epub 20190621. doi: 10.1038/s41467-019-10765-1. PubMed PMID: 31227709; PMCID: PMC6588589.

30. Rexrode LE, Hartley J, Showmaker KC, Challagundla L, Vandewege MW, Martin BE, Blair E, Bollavarapu R, Antonyraj RB, Hilton K, Gardiner A, Valeri J, Gisabella B, Garrett MR, Theoharides TC, Pantazopoulos H. Molecular profiling of the hippocampus of children with autism spectrum disorder. Mol Psychiatry. 2024. Epub 2024/02/15. doi: 10.1038/s41380-024-02441-8. PubMed PMID: 38355786.

31. Gogolla N, Caroni P, Luthi A, Herry C. Perineuronal nets protect fear memories from erasure. Science. 2009;325(5945):1258–61. Epub 2009/09/05. doi: 10.1126/science.1174146. PubMed PMID: 19729657.

32. Maeda N. Proteoglycans and neuronal migration in the cerebral cortex during development and disease. Frontiers in neuroscience. 2015;9:98. Epub 2015/04/09. doi: 10.3389/fnins.2015.00098. PubMed PMID: 25852466; PMCID: PMC4369650.

33. Karus M, Ulc A, Ehrlich M, Czopka T, Hennen E, Fischer J, Mizhorova M, Qamar N, Brustle O, Faissner A. Regulation of oligodendrocyte precursor maintenance by chondroitin sulphate glycosaminoglycans. Glia. 2016;64(2):270–86. Epub 2015/10/11. doi: 10.1002/glia.22928. PubMed PMID: 26454153.

34. Pomin VH. Sulfated glycans in inflammation. European journal of medicinal chemistry. 2015;92:353–69. Epub 2015/01/13. doi: 10.1016/j.ejmech.2015.01.002. PubMed PMID: 25576741.

35. Slaker ML, Jorgensen ET, Hegarty DM, Liu X, Kong Y, Zhang F, Linhardt RJ, Brown TE, Aicher SA, Sorg BA. Cocaine Exposure Modulates Perineuronal Nets and Synaptic Excitability of Fast-Spiking Interneurons in the Medial Prefrontal Cortex. eNeuro. 2018;5(5). Epub 2018/10/09. doi: 10.1523/ENEURO.0221-18.2018. PubMed PMID: 30294670; PMCID: PMC6171740.

36. Macke EL, Henningsen E, Jessen E, Zumwalde NA, Landowski M, Western DE, Lee WH, Liu C, Gruenke NP, Doebley AL, Miller S, Pattnaik B, Ikeda S, Gumperz JE, Ikeda A. Loss of Chondroitin Sulfate Modification Causes Inflammation and Neurodegeneration in skt Mice. Genetics. 2020;214(1):121–34. Epub 2019/11/23. doi: 10.1534/genetics.119.302834. PubMed PMID: 31754016; PMCID: PMC6944401.

37. Pitkanen A, Pikkarainen M, Nurminen N, Ylinen A. Reciprocal connections between the amygdala and the hippocampal formation, perirhinal cortex, and postrhinal cortex in rat. A review. Ann N Y Acad Sci. 2000;911:369–91. Epub 2000/07/27. doi: 10.1111/j.1749-6632.2000.tb06738.x. PubMed PMID: 10911886.

38. Benes FM, Berretta S. Amygdalo-entorhinal inputs to the hippocampal formation in relation to schizophrenia. Ann N Y Acad Sci. 2000;911:293–304. Epub 2000/07/27. doi: 10.1111/j.1749-6632.2000.tb06733.x. PubMed PMID: 10911881.

39. Bocchio M, Nabavi S, Capogna M. Synaptic Plasticity, Engrams, and Network Oscillations in Amygdala Circuits for Storage and Retrieval of Emotional Memories. Neuron. 2017;94(4):731–43. Epub 2017/05/19. doi: 10.1016/j.neuron.2017.03.022. PubMed PMID: 28521127.

40. Kasari C, Gulsrud AC, Wong C, Kwon S, Locke J. Randomized controlled caregiver mediated joint engagement intervention for toddlers with autism. Journal of autism and developmental disorders. 2010;40(9):1045–56. doi: 10.1007/s10803-010-0955-5. PubMed PMID: 20145986; PMCID: PMC2922697.

41. Dawson G, Rogers S, Munson J, Smith M, Winter J, Greenson J, Donaldson A, Varley J. Randomized, controlled trial of an intervention for toddlers with autism: the Early Start Denver Model. Pediatrics. 2010;125(1):e17–23. Epub 20091130. doi: 10.1542/peds.2009-0958. PubMed PMID: 19948568; PMCID: PMC4951085.

42. Tsilioni I, Patel AB, Pantazopoulos H, Berretta S, Conti P, Leeman SE, Theoharides TC. IL-37 is increased in brains of children with autism spectrum disorder and inhibits human microglia stimulated by neurotensin. Proc Natl Acad Sci U S A. 2019;116(43):21659–65. Epub 2019/10/09. doi: 10.1073/pnas.1906817116. PubMed PMID: 31591201; PMCID: PMC6815178.

43. Loomes R, Hull L, Mandy WPL. What Is the Male-to-Female Ratio in Autism Spectrum Disorder? A Systematic Review and Meta-Analysis. Journal of the American Academy of Child and Adolescent Psychiatry. 2017;56(6):466–74. Epub 2017/05/27. doi: 10.1016/j.jaac.2017.03.013. PubMed PMID: 28545751.

44. Jung M, Ma Y, Iyer RP, DeLeon-Pennell KY, Yabluchanskiy A, Garrett MR, Lindsey ML. IL-10 improves cardiac remodeling after myocardial infarction by stimulating M2 macrophage polarization and fibroblast activation. Basic research in cardiology. 2017;112(3):33. Epub 2017/04/26. doi: 10.1007/s00395-017-0622-5. PubMed PMID: 28439731; PMCID: PMC5575998.

45. Subramanian A, Tamayo P, Mootha VK, Mukherjee S, Ebert BL, Gillette MA, Paulovich A, Pomeroy SL, Golub TR, Lander ES, Mesirov JP. Gene set enrichment analysis: a knowledge-based approach for interpreting genome-wide expression profiles. Proc Natl Acad Sci U S A. 2005;102(43):15545–50. Epub 20050930. doi: 10.1073/pnas.0506580102. PubMed PMID: 16199517; PMCID: PMC1239896.

46. Tan F, Yang R, Xu X, Chen X, Wang Y, Ma H, Liu X, Wu X, Chen Y, Liu L, Jia X. Drug repositioning by applying ‘expression profiles’ generated by integrating chemical structure similarity and gene semantic similarity. Mol Biosyst. 2014;10(5):1126–38. Epub 2014/03/08. doi: 10.1039/c3mb70554d. PubMed PMID: 24603772.

47. Zaslavsky K, Zhang WB, McCready FP, Rodrigues DC, Deneault E, Loo C, Zhao M, Ross PJ, El Hajjar J, Romm A, Thompson T, Piekna A, Wei W, Wang Z, Khattak S, Mufteev M, Pasceri P, Scherer SW, Salter MW, Ellis J. SHANK2 mutations associated with autism spectrum disorder cause hyperconnectivity of human neurons. Nat Neurosci. 2019;22(4):556–64. Epub 20190325. doi: 10.1038/s41593-019-0365-8. PubMed PMID: 30911184; PMCID: PMC6475597.

48. Bi C, Wu J, Jiang T, Liu Q, Cai W, Yu P, Cai T, Zhao M, Jiang YH, Sun ZS. Mutations of ANK3 identified by exome sequencing are associated with autism susceptibility. Human mutation. 2012;33(12):1635–8. Epub 20120824. doi: 10.1002/humu.22174. PubMed PMID: 22865819.

49. Ashwood P, Enstrom A, Krakowiak P, Hertz-Picciotto I, Hansen RL, Croen LA, Ozonoff S, Pessah IN, Van de Water J. Decreased transforming growth factor beta1 in autism: a potential link between immune dysregulation and impairment in clinical behavioral outcomes. Journal of neuroimmunology. 2008;204(1-2):149–53. doi: 10.1016/j.jneuroim.2008.07.006. PubMed PMID: 18762342; PMCID: PMC2615583.

50. Ylisaukko-oja T, Rehnstrom K, Auranen M, Vanhala R, Alen R, Kempas E, Ellonen P, Turunen JA, Makkonen I, Riikonen R, Nieminen-von Wendt T, von Wendt L, Peltonen L, Jarvela I. Analysis of four neuroligin genes as candidates for autism. European journal of human genetics: EJHG. 2005;13(12):1285– 92. doi: 10.1038/sj.ejhg.5201474. PubMed PMID: 16077734.

51. Sudhof TC. Neuroligins and neurexins link synaptic function to cognitive disease. Nature. 2008;455(7215):903–11. doi: 10.1038/nature07456. PubMed PMID: 18923512; PMCID: PMC2673233.

52. Autism Genome Project C, Szatmari P, Paterson AD, Zwaigenbaum L, Roberts W, Brian J, Liu XQ, Vincent JB, Skaug JL, Thompson AP, Senman L, Feuk L, Qian C, Bryson SE, Jones MB, Marshall CR, Scherer SW, Vieland VJ, Bartlett C, Mangin LV, Goedken R, Segre A, Pericak-Vance MA, Cuccaro ML, Gilbert JR, Wright HH, Abramson RK, Betancur C, Bourgeron T, Gillberg C, Leboyer M, Buxbaum JD, Davis KL, Hollander E, Silverman JM, Hallmayer J, Lotspeich L, Sutcliffe JS, Haines JL, Folstein SE, Piven J, Wassink TH, Sheffield V, Geschwind DH, Bucan M, Brown WT, Cantor RM, Constantino JN, Gilliam TC, Herbert M, Lajonchere C, Ledbetter DH, Lese-Martin C, Miller J, Nelson S, Samango-Sprouse CA, Spence S, State M, Tanzi RE, Coon H, Dawson G, Devlin B, Estes A, Flodman P, Klei L, McMahon WM, Minshew N, Munson J, Korvatska E, Rodier PM, Schellenberg GD, Smith M, Spence MA, Stodgell C, Tepper PG, Wijsman EM, Yu CE, Roge B, Mantoulan C, Wittemeyer K, Poustka A, Felder B, Klauck SM, Schuster C, Poustka F, Bolte S, Feineis-Matthews S, Herbrecht E, Schmotzer G, Tsiantis J, Papanikolaou K, Maestrini E, Bacchelli E, Blasi F, Carone S, Toma C, Van Engeland H, de Jonge M, Kemner C, Koop F, Langemeijer M, Hijmans C, Staal WG, Baird G, Bolton PF, Rutter ML, Weisblatt E, Green J, Aldred C, Wilkinson JA, Pickles A, Le Couteur A, Berney T, McConachie H, Bailey AJ, Francis K, Honeyman G, Hutchinson A, Parr JR, Wallace S, Monaco AP, Barnby G, Kobayashi K, Lamb JA, Sousa I, Sykes N, Cook EH, Guter SJ, Leventhal BL, Salt J, Lord C, Corsello C, Hus V, Weeks DE, Volkmar F, Tauber M, Fombonne E, Shih A, Meyer KJ. Mapping autism risk loci using genetic linkage and chromosomal rearrangements. Nat Genet. 2007;39(3):319–28. Epub 20070218. doi: 10.1038/ng1985. PubMed PMID: 17322880; PMCID: PMC4867008.

53. Velmeshev D, Schirmer L, Jung D, Haeussler M, Perez Y, Mayer S, Bhaduri A, Goyal N, Rowitch DH, Kriegstein AR. Single-cell genomics identifies cell type-specific molecular changes in autism. Science. 2019;364(6441):685–9. Epub 2019/05/18. doi: 10.1126/science.aav8130. PubMed PMID: 31097668.

54. Hanamsagar R, Alter MD, Block CS, Sullivan H, Bolton JL, Bilbo SD. Generation of a microglial developmental index in mice and in humans reveals a sex difference in maturation and immune reactivity. Glia. 2018;66(2):460. Epub 2017/12/13. doi: 10.1002/glia.23277. PubMed PMID: 29230901.

55. Carlezon WA, Jr., Kim W, Missig G, Finger BC, Landino SM, Alexander AJ, Mokler EL, Robbins JO, Li Y, Bolshakov VY, McDougle CJ, Kim KS. Maternal and early postnatal immune activation produce sex-specific effects on autism-like behaviors and neuroimmune function in mice. Scientific reports. 2019;9(1):16928. Epub 2019/11/16. doi: 10.1038/s41598-019-53294-z. PubMed PMID: 31729416; PMCID: PMC6858355.

56. Haida O, Al Sagheer T, Balbous A, Francheteau M, Matas E, Soria F, Fernagut PO, Jaber M. Sex-dependent behavioral deficits and neuropathology in a maternal immune activation model of autism. Transl Psychiatry. 2019;9(1):124. Epub 2019/03/30. doi: 10.1038/s41398-019-0457-y. PubMed PMID: 30923308; PMCID: PMC6438965.

57. Dantzer R. Neuroimmune Interactions: From the Brain to the Immune System and Vice Versa. Physiol Rev. 2018;98(1):477–504. Epub 2018/01/20. doi: 10.1152/physrev.00039.2016. PubMed PMID: 29351513; PMCID: PMC5866360.

58. Glasson EJ, Bower C, Petterson B, de Klerk N, Chaney G, Hallmayer JF. Perinatal factors and the development of autism: a population study. Arch Gen Psychiatry. 2004;61(6):618–27. Epub 2004/06/09. doi: 10.1001/archpsyc.61.6.618. PubMed PMID: 15184241.

59. Gardener H, Spiegelman D, Buka SL. Perinatal and neonatal risk factors for autism: a comprehensive meta-analysis. Pediatrics. 2011;128(2):344–55. Epub 2011/07/13. doi: 10.1542/peds.2010-1036. PubMed PMID: 21746727; PMCID: PMC3387855.

60. Rosenberg GA, Cunningham LA, Wallace J, Alexander S, Estrada EY, Grossetete M, Razhagi A, Miller K, Gearing A. Immunohistochemistry of matrix metalloproteinases in reperfusion injury to rat brain: activation of MMP-9 linked to stromelysin-1 and microglia in cell cultures. Brain Res. 2001;893(1-2):104– 12. Epub 2001/02/27. doi: 10.1016/s0006-8993(00)03294-7. PubMed PMID: 11222998.

61. Cahoy JD, Emery B, Kaushal A, Foo LC, Zamanian JL, Christopherson KS, Xing Y, Lubischer JL, Krieg PA, Krupenko SA, Thompson WJ, Barres BA. A transcriptome database for astrocytes, neurons, and oligodendrocytes: a new resource for understanding brain development and function. J Neurosci. 2008;28(1):264–78. Epub 2008/01/04. doi: 10.1523/JNEUROSCI.4178-07.2008. PubMed PMID: 18171944; PMCID: PMC6671143.

62. Strackeljan L, Baczynska E, Cangalaya C, Baidoe-Ansah D, Wlodarczyk J, Kaushik R, Dityatev A. Microglia Depletion-Induced Remodeling of Extracellular Matrix and Excitatory Synapses in the Hippocampus of Adult Mice. Cells. 2021;10(8). Epub 20210722. doi: 10.3390/cells10081862. PubMed PMID: 34440631; PMCID: PMC8393852.

63. Thomas AL, Lehn MA, Janssen EM, Hildeman DA, Chougnet CA. Naturally-aged microglia exhibit phagocytic dysfunction accompanied by gene expression changes reflective of underlying neurologic disease. Scientific reports. 2022;12(1):19471. Epub 20221114. doi: 10.1038/s41598-022-21920-y. PubMed PMID: 36376530; PMCID: PMC9663419.

64. Ma D, Salyakina D, Jaworski JM, Konidari I, Whitehead PL, Andersen AN, Hoffman JD, Slifer SH, Hedges DJ, Cukier HN, Griswold AJ, McCauley JL, Beecham GW, Wright HH, Abramson RK, Martin ER, Hussman JP, Gilbert JR, Cuccaro ML, Haines JL, Pericak-Vance MA. A genome-wide association study of autism reveals a common novel risk locus at 5p14.1. Annals of human genetics. 2009;73(Pt 3):263–73. Epub 2009/05/22. doi: 10.1111/j.1469-1809.2009.00523.x. PubMed PMID: 19456320; PMCID: PMC2918410.

65. Wang K, Zhang H, Ma D, Bucan M, Glessner JT, Abrahams BS, Salyakina D, Imielinski M, Bradfield JP, Sleiman PM, Kim CE, Hou C, Frackelton E, Chiavacci R, Takahashi N, Sakurai T, Rappaport E, Lajonchere CM, Munson J, Estes A, Korvatska O, Piven J, Sonnenblick LI, Alvarez Retuerto AI, Herman EI, Dong H, Hutman T, Sigman M, Ozonoff S, Klin A, Owley T, Sweeney JA, Brune CW, Cantor RM, Bernier R, Gilbert JR, Cuccaro ML, McMahon WM, Miller J, State MW, Wassink TH, Coon H, Levy SE, Schultz RT, Nurnberger JI, Haines JL, Sutcliffe JS, Cook EH, Minshew NJ, Buxbaum JD, Dawson G, Grant SF, Geschwind DH, Pericak-Vance MA, Schellenberg GD, Hakonarson H. Common genetic variants on 5p14.1 associate with autism spectrum disorders. Nature. 2009;459(7246):528–33. Epub 2009/05/01. doi: 10.1038/nature07999. PubMed PMID: 19404256; PMCID: PMC2943511.

66. Weiss LA, Arking DE, Gene Discovery Project of Johns H, the Autism C, Daly MJ, Chakravarti A. A genome-wide linkage and association scan reveals novel loci for autism. Nature. 2009;461(7265):802–8. Epub 2009/10/09. doi: 10.1038/nature08490. PubMed PMID: 19812673; PMCID: PMC2772655.

67. Hussman JP, Chung RH, Griswold AJ, Jaworski JM, Salyakina D, Ma D, Konidari I, Whitehead PL, Vance JM, Martin ER, Cuccaro ML, Gilbert JR, Haines JL, Pericak-Vance MA. A noise-reduction GWAS analysis implicates altered regulation of neurite outgrowth and guidance in autism. Mol Autism. 2011;2(1):1. Epub 2011/01/21. doi: 10.1186/2040-2392-2-1. PubMed PMID: 21247446; PMCID: PMC3035032.

68. Anney R, Klei L, Pinto D, Regan R, Conroy J, Magalhaes TR, Correia C, Abrahams BS, Sykes N, Pagnamenta AT, Almeida J, Bacchelli E, Bailey AJ, Baird G, Battaglia A, Berney T, Bolshakova N, Bolte S, Bolton PF, Bourgeron T, Brennan S, Brian J, Carson AR, Casallo G, Casey J, Chu SH, Cochrane L, Corsello C, Crawford EL, Crossett A, Dawson G, de Jonge M, Delorme R, Drmic I, Duketis E, Duque F, Estes A, Farrar P, Fernandez BA, Folstein SE, Fombonne E, Freitag CM, Gilbert J, Gillberg C, Glessner JT, Goldberg J, Green J, Guter SJ, Hakonarson H, Heron EA, Hill M, Holt R, Howe JL, Hughes G, Hus V, Igliozzi R, Kim C, Klauck SM, Kolevzon A, Korvatska O, Kustanovich V, Lajonchere CM, Lamb JA, Laskawiec M, Leboyer M, Le Couteur A, Leventhal BL, Lionel AC, Liu XQ, Lord C, Lotspeich L, Lund SC, Maestrini E, Mahoney W, Mantoulan C, Marshall CR, McConachie H, McDougle CJ, McGrath J, McMahon WM, Melhem NM, Merikangas A, Migita O, Minshew NJ, Mirza GK, Munson J, Nelson SF, Noakes C, Noor A, Nygren G, Oliveira G, Papanikolaou K, Parr JR, Parrini B, Paton T, Pickles A, Piven J, Posey DJ, Poustka A, Poustka F, Prasad A, Ragoussis J, Renshaw K, Rickaby J, Roberts W, Roeder K, Roge B, Rutter ML, Bierut LJ, Rice JP, Salt J, Sansom K, Sato D, Segurado R, Senman L, Shah N, Sheffield VC, Soorya L, Sousa I, Stoppioni V, Strawbridge C, Tancredi R, Tansey K, Thiruvahindrapduram B, Thompson AP, Thomson S, Tryfon A, Tsiantis J, Van Engeland H, Vincent JB, Volkmar F, Wallace S, Wang K, Wang Z, Wassink TH, Wing K, Wittemeyer K, Wood S, Yaspan BL, Zurawiecki D, Zwaigenbaum L, Betancur C, Buxbaum JD, Cantor RM, Cook EH, Coon H, Cuccaro ML, Gallagher L, Geschwind DH, Gill M, Haines JL, Miller J, Monaco AP, Nurnberger JI, Jr., Paterson AD, Pericak-Vance MA, Schellenberg GD, Scherer SW, Sutcliffe JS, Szatmari P, Vicente AM, Vieland VJ, Wijsman EM, Devlin B, Ennis S, Hallmayer J. A genome-wide scan for common alleles affecting risk for autism. Hum Mol Genet. 2010;19(20):4072–82. Epub 2010/07/29. doi: 10.1093/hmg/ddq307. PubMed PMID: 20663923; PMCID: PMC2947401.

69. Bilousova TV, Dansie L, Ngo M, Aye J, Charles JR, Ethell DW, Ethell IM. Minocycline promotes dendritic spine maturation and improves behavioural performance in the fragile X mouse model. Journal of medical genetics. 2009;46(2):94–102. Epub 2008/10/07. doi: 10.1136/jmg.2008.061796. PubMed PMID: 18835858.

70. Wen TH, Afroz S, Reinhard SM, Palacios AR, Tapia K, Binder DK, Razak KA, Ethell IM. Genetic Reduction of Matrix Metalloproteinase-9 Promotes Formation of Perineuronal Nets Around Parvalbumin-Expressing Interneurons and Normalizes Auditory Cortex Responses in Developing Fmr1 Knock-Out Mice. Cereb Cortex. 2018;28(11):3951–64. Epub 2017/10/19. doi: 10.1093/cercor/bhx258. PubMed PMID: 29040407; PMCID: PMC6188540.

71. Pirbhoy PS, Rais M, Lovelace JW, Woodard W, Razak KA, Binder DK, Ethell IM. Acute pharmacological inhibition of matrix metalloproteinase-9 activity during development restores perineuronal net formation and normalizes auditory processing in Fmr1 KO mice. J Neurochem. 2020. Epub 2020/05/07. doi: 10.1111/jnc.15037. PubMed PMID: 32374912.

72. Reinhard SM, Rais M, Afroz S, Hanania Y, Pendi K, Espinoza K, Rosenthal R, Binder DK, Ethell IM, Razak KA. Reduced perineuronal net expression in Fmr1 KO mice auditory cortex and amygdala is linked to impaired fear-associated memory. Neurobiol Learn Mem. 2019;164:107042. Epub 2019/07/22. doi: 10.1016/j.nlm.2019.107042. PubMed PMID: 31326533.

73. Miyata S, Kitagawa H. Formation and remodeling of the brain extracellular matrix in neural plasticity: Roles of chondroitin sulfate and hyaluronan. Biochim Biophys Acta Gen Subj. 2017;1861(10):2420–34. Epub 20170615. doi: 10.1016/j.bbagen.2017.06.010. PubMed PMID: 28625420.

74. Levy AD, Omar MH, Koleske AJ. Extracellular matrix control of dendritic spine and synapse structure and plasticity in adulthood. Front Neuroanat. 2014;8:116. Epub 20141020. doi: 10.3389/fnana.2014.00116. PubMed PMID: 25368556; PMCID: PMC4202714.

75. Ferrer-Ferrer M, Dityatev A. Shaping Synapses by the Neural Extracellular Matrix. Front Neuroanat. 2018;12:40. Epub 20180515. doi: 10.3389/fnana.2018.00040. PubMed PMID: 29867379; PMCID: PMC5962695.

76. Singh JB, Perello-Amoros B, Schneeberg J, Mirzapourdelavar H, Seidenbecher CI, Fejtova A, Dityatev A, Frischknecht R. Activity-dependent extracellular proteolytic cascade cleaves the ECM component brevican to promote structural plasticity. EMBO reports. 2026;27(1):163–85. Epub 20251119. doi: 10.1038/s44319-025-00644-w. PubMed PMID: 41261283; PMCID: PMC12796228.

77. Tabet A, Apra C, Stranahan AM, Anikeeva P. Changes in Brain Neuroimmunology Following Injury and Disease. Frontiers in integrative neuroscience. 2022;16:894500. Epub 2022/05/17. doi: 10.3389/fnint.2022.894500. PubMed PMID: 35573444; PMCID: PMC9093707.

78. Coll-Tane M, Gong NN, Belfer SJ, van Renssen LV, Kurtz-Nelson EC, Szuperak M, Eidhof I, van Reijmersdal B, Terwindt I, Durkin J, Verheij MMM, Kim CN, Hudac CM, Nowakowski TJ, Bernier RA, Pillen S, Earl RK, Eichler EE, Kleefstra T, Kayser MS, Schenck A. The CHD8/CHD7/Kismet family links blood-brain barrier glia and serotonin to ASD-associated sleep defects. Sci Adv. 2021;7(23). Epub 2021/06/06. doi: 10.1126/sciadv.abe2626. PubMed PMID: 34088660; PMCID: PMC8177706.

79. Fiorentino M, Sapone A, Senger S, Camhi SS, Kadzielski SM, Buie TM, Kelly DL, Cascella N, Fasano A. Blood-brain barrier and intestinal epithelial barrier alterations in autism spectrum disorders. Mol Autism. 2016;7:49. Epub 2016/12/14. doi: 10.1186/s13229-016-0110-z. PubMed PMID: 27957319; PMCID: PMC5129651.

80. Li H, Wang X, Hu C, Li H, Xu Z, Lei P, Luo X, Hao Y. JUN and PDGFRA as Crucial Candidate Genes for Childhood Autism Spectrum Disorder. Front Neuroinform. 2022;16:800079. Epub 20220516. doi: 10.3389/fninf.2022.800079. PubMed PMID: 35655651; PMCID: PMC9152672.

81. Kajizuka M, Miyachi T, Matsuzaki H, Iwata K, Shinmura C, Suzuki K, Suda S, Tsuchiya KJ, Matsumoto K, Iwata Y, Nakamura K, Tsujii M, Sugiyama T, Takei N, Mori N. Serum levels of platelet-derived growth factor BB homodimers are increased in male children with autism. Prog Neuropsychopharmacol Biol Psychiatry. 2010;34(1):154–8. Epub 20091029. doi: 10.1016/j.pnpbp.2009.10.017. PubMed PMID: 19879307.

82. Xiang C, Li H, Tang W. Targeting CSF-1R represents an effective strategy in modulating inflammatory diseases. Pharmacol Res. 2023;187:106566. Epub 20221121. doi: 10.1016/j.phrs.2022.106566. PubMed PMID: 36423789.

83. Irvine KM, Burns CJ, Wilks AF, Su S, Hume DA, Sweet MJ. A CSF-1 receptor kinase inhibitor targets effector functions and inhibits pro-inflammatory cytokine production from murine macrophage populations. Faseb J. 2006;20(11):1921–3. Epub 20060728. doi: 10.1096/fj.06-5848fje. PubMed PMID: 16877523.

84. Kane JL, Jr., Asmussen G, Batchelor J, Cromwell M, Fezoui M, Fitzgerald M, Giese B, Gladysheva T, Holley S, Keefe K, Kothe M, Lam B, Lim S, Liu J, Ma L, Metz M, Scholte AA, Shum P, Wei L, Woodworth L, Edling A. Identification of Selective Imidazopyridine CSF1R Inhibitors. ACS Med Chem Lett. 2024;15(5):722–30. Epub 20240430. doi: 10.1021/acsmedchemlett.4c00110. PubMed PMID: 38746878; PMCID: PMC11089549.

85. Erbas O, Solmaz V, Aksoy D. Inhibitor effect of dexketoprofen in rat model of pentylenetetrazol-induced seizures. Neurol Res. 2015;37(12):1096–101. Epub 20140526. doi: 10.1179/1743132814Y.0000000391. PubMed PMID: 24861495.

86. Alvino FG, Gini S, Minetti A, Pagani M, Sastre-Yague D, Barsotti N, De Guzman E, Schleifer C, Stuefer A, Kushan L, Montani C, Galbusera A, Papaleo F, Kates WR, Murphy D, Lombardo MV, Pasqualetti M, Bearden CE, Gozzi A. Synaptic-dependent developmental dysconnectivity in 22q11.2 deletion syndrome. Sci Adv. 2025;11(11):eadq2807. Epub 20250312. doi: 10.1126/sciadv.adq2807. PubMed PMID: 40073125; PMCID: PMC11900866.

87. McCamphill PK, Stoppel LJ, Senter RK, Lewis MC, Heynen AJ, Stoppel DC, Sridhar V, Collins KA, Shi X, Pan JQ, Madison J, Cottrell JR, Huber KM, Scolnick EM, Holson EB, Wagner FF, Bear MF. Selective inhibition of glycogen synthase kinase 3alpha corrects pathophysiology in a mouse model of fragile X syndrome. Science translational medicine. 2020;12(544). doi: 10.1126/scitranslmed.aam8572. PubMed PMID: 32434848; PMCID: PMC8095719.

88. Rizk M, Saker Z, Harati H, Fares Y, Bahmad HF, Nabha S. Deciphering the roles of glycogen synthase kinase 3 (GSK3) in the treatment of autism spectrum disorder and related syndromes. Molecular biology reports. 2021;48(3):2669–86. Epub 20210301. doi: 10.1007/s11033-021-06237-9. PubMed PMID: 33650079.

89. Madden JM, Lakoma MD, Lynch FL, Rusinak D, Owen-Smith AA, Coleman KJ, Quinn VP, Yau VM, Qian YX, Croen LA. Psychotropic Medication Use among Insured Children with Autism Spectrum Disorder. Journal of autism and developmental disorders. 2017;47(1):144–54. doi: 10.1007/s10803-016-2946-7. PubMed PMID: 27817163; PMCID: PMC5328123.

90. Coury DL, Anagnostou E, Manning-Courtney P, Reynolds A, Cole L, McCoy R, Whitaker A, Perrin JM. Use of psychotropic medication in children and adolescents with autism spectrum disorders. Pediatrics. 2012;130 Suppl 2:S69–76. doi: 10.1542/peds.2012-0900D. PubMed PMID: 23118256.

91. Oswald DP, Sonenklar NA. Medication use among children with autism spectrum disorders. Journal of child and adolescent psychopharmacology. 2007;17(3):348–55. doi: 10.1089/cap.2006.17303. PubMed PMID: 17630868.

92. Houghton R, Ong RC, Bolognani F. Psychiatric comorbidities and use of psychotropic medications in people with autism spectrum disorder in the United States. Autism Res. 2017;10(12):2037–47. Epub 20170930. doi: 10.1002/aur.1848. PubMed PMID: 28963809.

93. Manter MA, Birtwell KB, Bath J, Friedman NDB, Keary CJ, Neumeyer AM, Palumbo ML, Thom RP, Stonestreet E, Brooks H, Dakin K, Hooker JM, McDougle CJ. Pharmacological treatment in autism: a proposal for guidelines on common co-occurring psychiatric symptoms. BMC medicine. 2025;23(1):11. Epub 20250107. doi: 10.1186/s12916-024-03814-0. PubMed PMID: 39773705; PMCID: PMC11705908.

94. Karthikeyan R, Cardinali DP, Shakunthala V, Spence DW, Brown GM, Pandi-Perumal SR. Understanding the role of sleep and its disturbances in Autism spectrum disorder. Int J Neurosci. 2020;130(10):1033–46. Epub 20200114. doi: 10.1080/00207454.2019.1711377. PubMed PMID: 31903819.

95. Liu X, Hubbard JA, Fabes RA, Adam JB. Sleep disturbances and correlates of children with autism spectrum disorders. Child Psychiatry Hum Dev. 2006;37(2):179–91. Epub 2006/09/27. doi: 10.1007/s10578-006-0028-3. PubMed PMID: 17001527.

96. Cohen S, Conduit R, Lockley SW, Rajaratnam SM, Cornish KM. The relationship between sleep and behavior in autism spectrum disorder (ASD): a review. Journal of neurodevelopmental disorders. 2014;6(1):44. Epub 2014/12/23. doi: 10.1186/1866-1955-6-44. PubMed PMID: 25530819; PMCID: PMC4271434.

97. Souders MC, Zavodny S, Eriksen W, Sinko R, Connell J, Kerns C, Schaaf R, Pinto-Martin J. Sleep in Children with Autism Spectrum Disorder. Curr Psychiatry Rep. 2017;19(6):34. Epub 2017/05/16. doi: 10.1007/s11920-017-0782-x. PubMed PMID: 28502070; PMCID: PMC5846201.

98. Smith AM, Johnson AH, Bashore L. Exploration of sleep disturbances in children and adolescents with and without autism in a paediatric sample referred for polysomnography. J Paediatr Child Health. 2023;59(8):948–54. Epub 2023/05/10. doi: 10.1111/jpc.16421. PubMed PMID: 37162017.

99. Ahmmad MR, Pantazopoulos H, Faruque F, Zhang X, Puri R. Association Between Age-Specific Sleep Sufficiency and Autism Spectrum Disorder in U.S. Children. Journal of autism and developmental disorders. 2025. Epub 20251212. doi: 10.1007/s10803-025-07175-2. PubMed PMID: 41385124.

100. Jones CE, Opel RA, Kaiser ME, Chau AQ, Quintana JR, Nipper MA, Finn DA, Hammock EAD, Lim MM. Early-life sleep disruption increases parvalbumin in primary somatosensory cortex and impairs social bonding in prairie voles. Sci Adv. 2019;5(1):eaav5188. Epub 2019/02/08. doi: 10.1126/sciadv.aav5188. PubMed PMID: 30729165; PMCID: PMC6353622.

101. Jones CE, Chau AQ, Olson RJ, Moore C, Wickham PT, Puranik N, Guizzetti M, Cao H, Meshul CK, Lim MM. Early life sleep disruption alters glutamate and dendritic spines in prefrontal cortex and impairs cognitive flexibility in prairie voles. Curr Res Neurobiol. 2021;2. Epub 2021/01/01. doi: 10.1016/j.crneur.2021.100020. PubMed PMID: 35505895; PMCID: PMC9060254.

102. Jones-Tinsley CE, Olson RJ, Mader M, Wickham PT, Gutowsky K, Wong C, Chu SS, Milman NEP, Cao H, Lim MM. Early life sleep disruption has long lasting, sex specific effects on later development of sleep in prairie voles. Neurobiol Sleep Circadian Rhythms. 2023;14:100087. Epub 2023/01/31. doi: 10.1016/j.nbscr.2022.100087. PubMed PMID: 36712905; PMCID: PMC9879777.

103. Yan T, Goldman RD. Melatonin for children with autism spectrum disorder. Canadian family physician Medecin de famille canadien. 2020;66(3):183–5. PubMed PMID: 32165465; PMCID: PMC8302336.

104. Gagnon K, Godbout R. Melatonin and Comorbidities in Children with Autism Spectrum Disorder. Curr Dev Disord Rep. 2018;5(3):197–206. Epub 20180809. doi: 10.1007/s40474-018-0147-0. PubMed PMID: 30148039; PMCID: PMC6096870.

105. Malow BA, Findling RL, Schroder CM, Maras A, Breddy J, Nir T, Zisapel N, Gringras P. Sleep, Growth, and Puberty After 2 Years of Prolonged-Release Melatonin in Children With Autism Spectrum Disorder. Journal of the American Academy of Child and Adolescent Psychiatry. 2021;60(2):252–61 e3. Epub 20200123. doi: 10.1016/j.jaac.2019.12.007. PubMed PMID: 31982581; PMCID: PMC8084705.

106. Wray JA, Yoon JH, Vollmer T, Mauk J. Pilot study of the behavioral effects of flumazenil in two children with autism. Journal of autism and developmental disorders. 2000;30(6):619–20. doi: 10.1023/a:1005651829910. PubMed PMID: 11261474.

107. Ming X, Gordon E, Kang N, Wagner GC. Use of clonidine in children with autism spectrum disorders. Brain & development. 2008;30(7):454–60. Epub 20080215. doi: 10.1016/j.braindev.2007.12.007. PubMed PMID: 18280681.

108. Jacobson LH, Hoyer D, de Lecea L. Hypocretins (orexins): The ultimate translational neuropeptides. J Intern Med. 2022;291(5):533–56. Epub 20220119. doi: 10.1111/joim.13406. PubMed PMID: 35043499.

109. Louros SR, Seo SS, Maio B, Martinez-Gonzalez C, Gonzalez-Lozano MA, Muscas M, Verity NC, Wills JC, Li KW, Nolan MF, Osterweil EK. Excessive proteostasis contributes to pathology in fragile X syndrome. Neuron. 2023;111(4):508–25 e7. Epub 20221209. doi: 10.1016/j.neuron.2022.11.012. PubMed PMID: 36495869.

110. Christensen J, Gronborg TK, Sorensen MJ, Schendel D, Parner ET, Pedersen LH, Vestergaard M. Prenatal valproate exposure and risk of autism spectrum disorders and childhood autism. Jama. 2013;309(16):1696–703. doi: 10.1001/jama.2013.2270. PubMed PMID: 23613074; PMCID: PMC4511955.

111. Wiggs KK, Rickert ME, Sujan AC, Quinn PD, Larsson H, Lichtenstein P, Oberg AS, D’Onofrio BM. Antiseizure medication use during pregnancy and risk of ASD and ADHD in children. Neurology. 2020;95(24):e3232–e40. Epub 20201028. doi: 10.1212/WNL.0000000000010993. PubMed PMID: 33115775; PMCID: PMC7836668.

112. Nicolini C, Fahnestock M. The valproic acid-induced rodent model of autism. Exp Neurol. 2018;299(Pt A):217–27. Epub 20170502. doi: 10.1016/j.expneurol.2017.04.017. PubMed PMID: 28472621.

